# Infectivity, susceptibility, and risk factors associated with SARS-CoV-2 transmission under intensive contact tracing in Hunan, China

**DOI:** 10.1101/2020.07.23.20160317

**Authors:** Shixiong Hu, Wei Wang, Yan Wang, Maria Litvinova, Kaiwei Luo, Lingshuang Ren, Qianlai Sun, Xinghui Chen, Ge Zeng, Jing Li, Lu Liang, Zhihong Deng, Wen Zheng, Mei Li, Hao Yang, Jinxin Guo, Kai Wang, Xinhua Chen, Ziyan Liu, Han Yan, Huilin Shi, Zhiyuan Chen, Yonghong Zhou, Kaiyuan Sun, Alessandro Vespignani, Cécile Viboud, Lidong Gao, Marco Ajelli, Hongjie Yu

**Author notes:** **Corresponding Authors:** Lidong Gao, MSc, Hunan Provincial Center for Disease Control and Prevention, No. 450 Middle Furong Road, Kaifu District, 410005, Changsha, China, Marco Ajelli, PhD, Department of Epidemiology and Biostatistics, Indiana University School of Public Health, Bloomington, 47405 IN, USA, Hongjie Yu, PhD, School of Public Health, Fudan University, Key Laboratory of Public Health Safety, Ministry of Education, No. 138 Yixueyuan Road, Xuhui District, 200032, Shanghai, China. These authors are joint first authors and contributed equally to this work (Shixiong Hu, Wei Wang, Yan Wang, Maria Litvinova). These authors are joint senior authors and contributed equally to this work (Lidong Gao, Marco Ajelli, Hongjie Yu).

## Abstract

Several mechanisms driving SARS-CoV-2 transmission remain unclear. Based on individual records of 1,178 SARS-CoV-2 infectors and their 15,648 contacts in Hunan, China, we estimated key transmission parameters. The mean generation time was estimated to be 5.7 (median: 5.5, IQR: 4.5, 6.8) days, with infectiousness peaking 1.8 days before symptom onset, with 95% of transmission events occurring between 8.8 days before and 9.5 days after symptom onset. Most of transmission events occurred during the pre-symptomatic phase (59.2%). SARS-CoV-2 susceptibility to infection increases with age, while transmissibility is not significantly different between age groups and between symptomatic and asymptomatic individuals. Contacts in households and exposure to first-generation cases are associated with higher odds of transmission. Our findings support the hypothesis that children can effectively transmit SARS-CoV-2 and highlight how pre-symptomatic and asymptomatic transmission can hinder control efforts.

## Introduction

The outbreak of coronavirus disease 2019 (COVID-19) started in December 2009 in Wuhan, China ^1^. The outbreak, caused by the SARS-CoV-2 virus, quickly spread globally, leading WHO to declare a pandemic on March 11, 2020 ^2^. Despite more than 18.4 million SARS-CoV-2 infected individuals confirmed worldwide as of August 06, 2020 ^3^, there are still many unknowns in the epidemiology and natural history of COVID-19.

A key question under debate is whether the infectivity of individuals with, and susceptibility to, SARS-CoV-2 infection differs by age. In particular, the role of children in SARS-CoV-2 transmission has yet to be fully understood. Schools were closed in the early months of the pandemic in most countries ^4,5^, so that the low proportion of cases notified in young individuals ^6^ could be attributed to a low probability of developing symptoms ^7,8^, a low susceptibility to infection ^9-11^, and/or few contact opportunities relative to other age groups. The importance of each of these factors has been difficult thus far to disentangle. A related question is the probability of asymptomatic transmission. In fact, it is often argued that the COVID-19 pandemic has been difficult to tackle because of the importance of pre-symptomatic and asymptomatic transmission. Evidence from confined settings such households, homeless shelters, and nursing facilities, supports the role of pre-symptomatic and asymptomatic transmission ^1,10,12-15^. Yet, a quantification of the contribution of asymptomatic and pre-symptomatic transmission in large populations is still lacking.

A full understanding of SARS-CoV-2 transmission patterns and risk factors is crucial to plan targeted COVID-19 responses, especially as countries relax costly lockdown policies and move towards case-based interventions (e.g., case isolation, quarantine of contacts, contact tracing). To define the temporal characteristics of the response strategies (e.g., duration of the quarantine and isolation period, definition of contacts to be traced) it is crucial to understand the age profile of infectiousness and to have robust estimates of key time-to-event distributions such as the generation time. These distributions were estimated in the early days of the pandemic based on the very first few clusters of cases and are thus subject to high uncertainty and variability between different studies ^1,16^. It is important to update these estimates using large-scale and harmonized epidemiological datasets.

In this study, we analyze 1,178 SARS-CoV-2 infected individuals and their 15,648 contacts identified by contact tracing operations carried out in the Hunan Province of China over the period from January 13-April 02, 2020. This comprehensive and detailed dataset compiled by the Hunan Provincial CDC sheds light on SARS-CoV-2 transmission risk factors, and the distribution of key time-to-event parameters.

## Results

### Sample description

Between January 23, 2020 and April 02, 2020, 1,019 symptomatic cases and 159 asymptomatic subjects were reported and screened for inclusion (Fig. S1 and Tab. 1). Through active contacts tracing, a total of 15,648 close contacts were identified, of whom 471 contacts were positive for SARS-CoV-2 infection. Among 1,178 SARS-CoV-2 infections, we identified an infector for a total of 432 transmission events, 831 epidemiologically linked cases (including index cases) in 210 clusters. Of these clusters, 499 SARS-CoV-2 infections in 123 clusters had a clear epidemiological link to a previous SARS-CoV-2 infected individual. From 15,648 close contacts, 6,412 were identified by forward contact tracing and resulted in the identification of 285 symptomatic cases and 63 asymptomatic SARS-CoV-2 positive subjects. The remaining 9,236 close contacts were identified through backward contact tracing. The distribution of the cases and close contacts in time and space is presented in Fig. 1 and Fig. S2-3. Overall, the median age of symptomatic cases and asymptomatic subjects, and their close contacts were 45 (IQR: 34-55), 36 (IQR: 19-52) and 40 (IQR: 27-52) years, respectively (Tab. 1). Cases aged 0-14 years presented milder or no clinical symptoms, while patients aged 15 years and older had more severe illness (P<0.001) (Fig. S4D).

**Figure 1.**
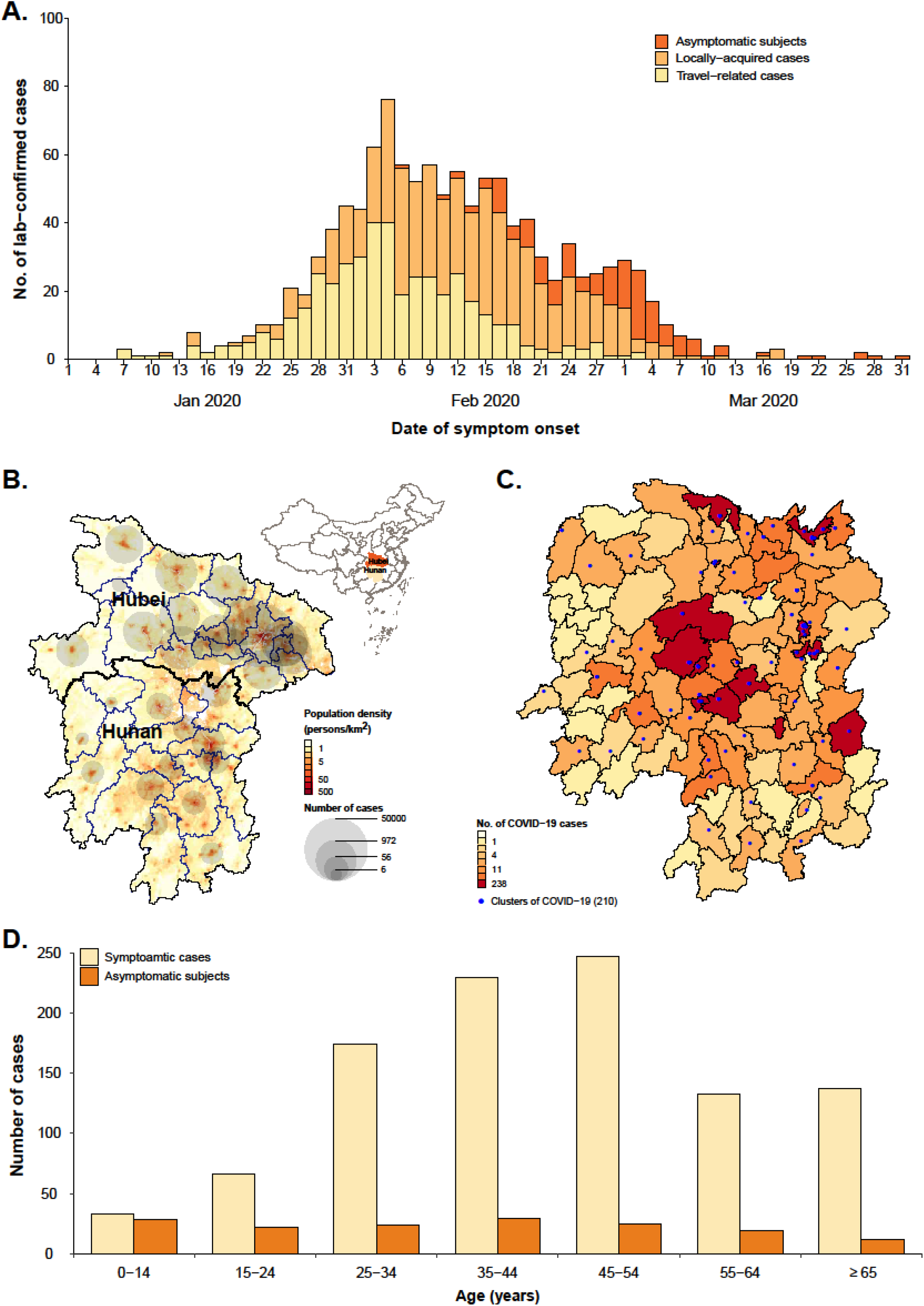
Temporal, geographical and age distribution of SARS-CoV-2 infected individuals stratified by the presence of symptoms and source of infection in Hunan Province, China. (A) Daily number of new SARS-CoV-2 infected individuals by date of symptom onset by source of infection For asymptomatic individuals we used date of the first RT-PCR positive result. (B) Geographical distribution of SARS-CoV-2 infected individuals in Hunan Province, and geo-locations of Hubei and Hunan provinces. (C) Geographical distribution of SARS-CoV-2 clusters. (D) Age distribution of SARS-CoV-2 symptomatic and asymptomatic infected individuals.

### Time-to-key-event distributions

To estimate the duration of the incubation period, we analyzed 268 locally-acquired infections belonging to 114 clusters, with information on both the date(s) of exposure and transmission generation in the cluster. We found that the best fitting distribution of the incubation period was a Weibull distribution with mean 6.4 days (median: 5.7, IQR: 3.2-8.8) (Tab. S3). We performed two sensitivity analyses, one excluding cases with only an exposure end date (17 individuals) and another one where we inferred the earliest exposure date for 7 of those 17 individuals. The results of both sensitivity analyses are consistent with the main analysis (Supporting Information, Tab. S3).

Symptom onset dates were available for 245 transmission pairs; the resulting serial interval had an estimated mean of 5.5 days (median: 4.8, IQR: 0.9-9.4), based on fitting a gamma distribution. By considering only pairs with a single identified infector, we find that 14.0% (31/221) of the empirical serial intervals were negative, which means that the symptom onset date of the infectee precedes the symptom onset date of her/his infector. To assess whether the serial interval changed over the course of the epidemic, we divided the outbreak in Hunan in two periods: the first one running from the detection of the first case up to January 23 and the second one from January 24 (date of the activation of the Level 1 Emergency Response) to April 2 (date of the detection of the last confirmed case). The mean serial interval decreased from 7.0 (median: 6.6, IQR: 3.5-10.1) days in the first period, to 4.1 (median: 3.2, IQR: −1.1, 8.4) days in the second period.

The mean generation time was estimated to be 5.7 days (median: 5.5, IQR: 4.5-6.8). The difference between the estimated mean incubation period and mean generation time is less than one day (Fig. 2B).

**Figure 2.**
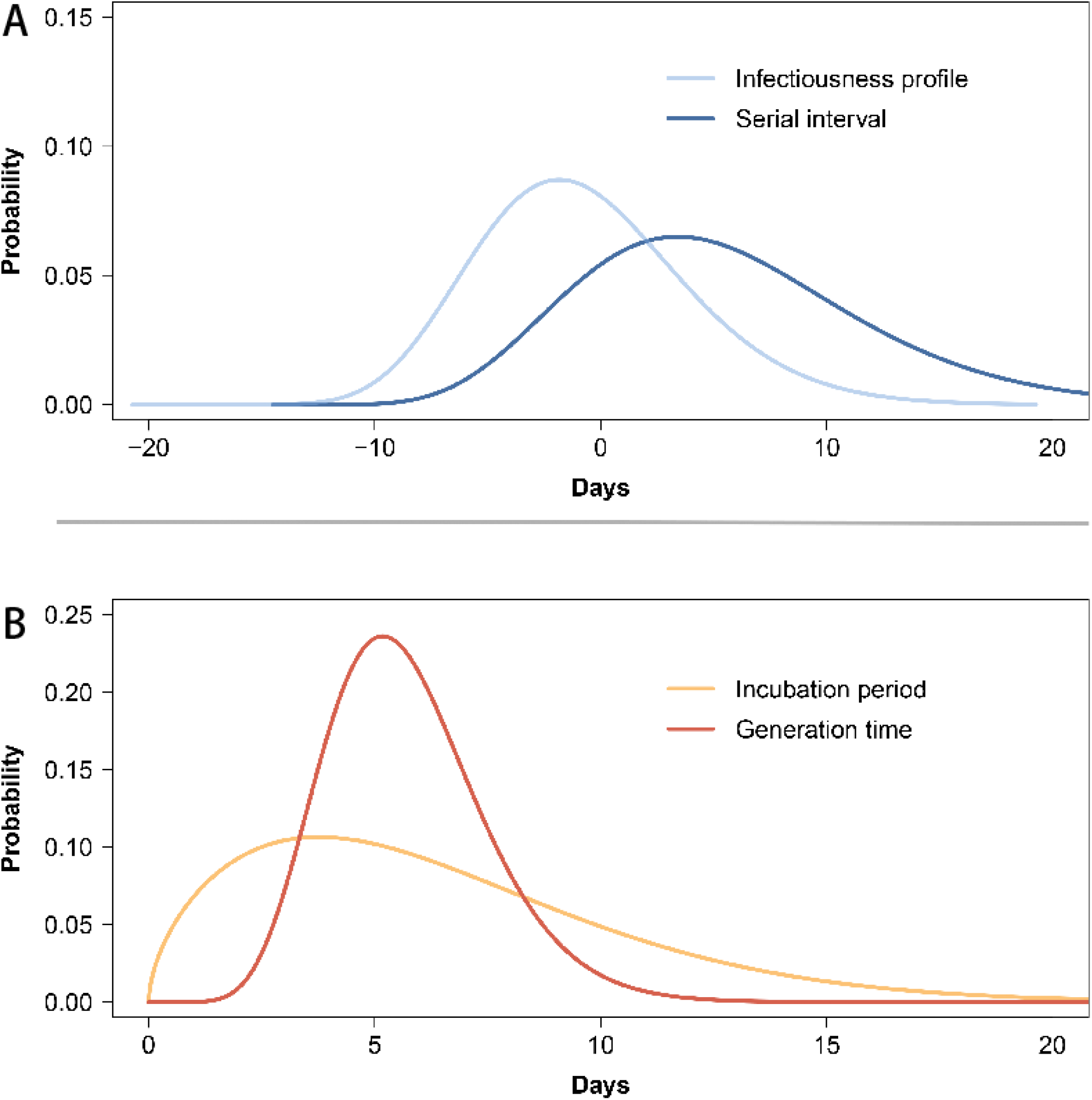
Quantifying the serial interval, infectiousness profile, incubation period and generation time fitted by gamma or Weibull distributions. (A) Estimated distribution of the serial interval and of the infectiousness profile by gamma distributions based on 245 transmission pairs. (B) Estimated distribution of the incubation period by Weibull distributions and of the generation time by gamma distributions based on 268 locally-acquired confirmed cases belonging to 114 clusters.

The mean time interval from symptom onset to the date of collection of the sample for PCR testing was estimated to be 4.7 days (median: 4.2, IQR: 1.4-7.4) using the best fitting gamma distribution, based on 531 confirmed cases. The mean time interval from symptom onset to laboratory confirmation was estimated to be 6.4 (median: 6.0, IQR:3.3-9.1) days, based on 952 confirmed cases.

### Pre-symptomatic transmission

Infectiousness was estimated to peak 1.8 days before symptom onset (Fig. 2A). It is important to stress that our estimate provides a measure of the probability of infecting contacts at any time after the time of exposure to the infector. As such, this is an empirical measure of the transmissibility over time of infectors, which accounts for human behavior (contacts) and performed interventions (e.g., case isolation, precautionary behaviors). We estimated the proportion of pre-symptomatic transmission (area under the curve, Fig. 2A) at 59.2%, with 95% of transmission events occurring between −8.8 days and 9.5 days of the date of symptom onset. The proportion of pre-symptomatic transmission (area under the curve) increased from 50.8% for the period from January 5 to January 23, to 76.7% for the period from January 24 to April 2, when intensive contact tracing and isolation strategy were undertaken by the Hunan Province. From the analysis of the transmission chains reconstructed by field investigations, 43 pre-symptomatic transmission events were recorded in 23 clusters. A subset of those clusters is shown in Fig. 3.

**Figure 3.**
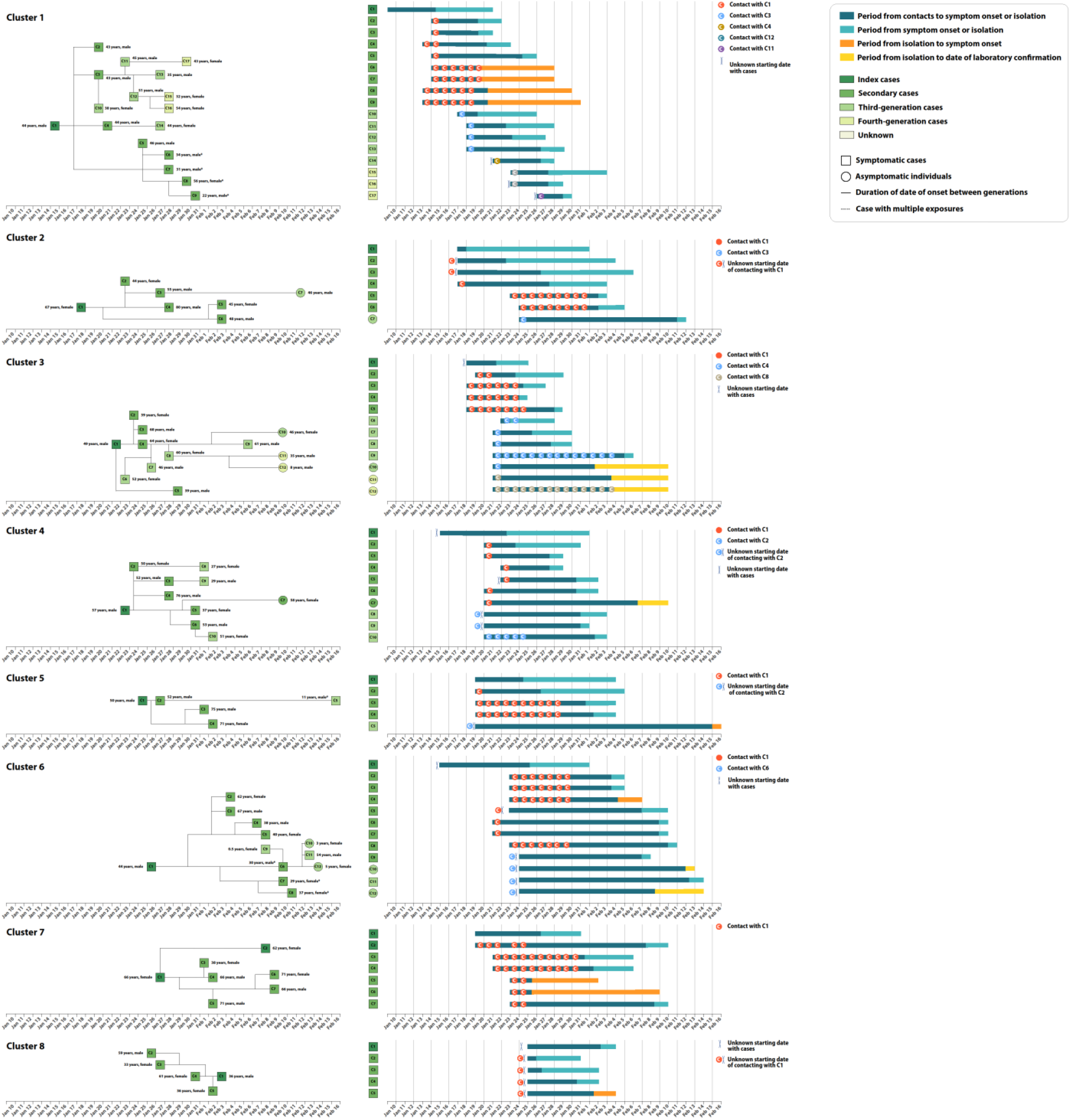
Timing of transmission events and SARS-CoV-2 infected individuals in randomly selected clusters showing evidence of pre-symptomatic transmission. Square symbols indicate symptomatic cases and circular symbols indicate asymptomatic subjects. Age, sex and generation in a cluster are shown for each SARS-CoV-2 infected individual (left panels), with information on date of illness onset for symptomatic cases and date of diagnosis to the first RT-PCR positive for asymptomatic subjects and individuals without date of illness onset (symbol “*”). Timeline of events (right panels).

### Asymptomatic transmission

From the analysis of contact tracing records, we identified 8 clusters (25 local transmission events) with evidence of asymptomatic transmission. There were 11 asymptomatic infectors (5 primary and 6 secondary infectors) associated with 15 transmission events (10 secondary and 5 tertiary infections, Fig. S5). No asymptomatic individual was a cluster index case (i.e., did not trigger a contact tracing investigation), although 5 of them were primary infectors.

### SARS-CoV-2 risk factors

We first explored differences in the age of SARS-CoV-2 infectors and infectees through the construction of age-specific transmission matrices (Fig. S6). The results suggest that people aged 15-64 years generated a larger mean number of cases than younger (0-14 years old) and older (65+ years old) individuals. Moreover, individuals aged 65 years and older were infected more often. Note that these three age groups were chosen to represent three key segments of the population, namely i) younger than working-age (students and preschoolers), ii) working-age population, and iii) retiring-age individuals. We have also examined the proportion of transmission events associated with asymptomatic infectors. In our sample, we had 432 transmission events with an identified infector, 15 of which (3.5%) were associated with asymptomatic infectors. However, the share increases to 8.5% (10/118) if we focus on transmission events occurring after February 7, 2020 and to 60% (15/25) if we consider only clusters with at least one asymptomatic transmission event.

It is important to stress that these estimates do not account for several confounding factors (e.g., all index cases are symptomatic, children are more likely to be in later generations of transmission, see Supporting Information Tab. S6 and S7). To account for the possible effect of multiple confounding factors, we thus performed a multivariate regression analysis (GLMM). We found that the age of the contact, the contact setting, and the generation of the infector in a cluster were important risk factors for transmission (Tab. 2). Infectiousness was not significantly different between working-age adults (15-64 years old) and other age groups (0-14 years old: p-value=0.210; 65 years and over: p-value=0.306); in contrast, susceptibility to SARS-CoV-2 infection increased with age (p-value=0.028, Model 2 in Tab. 2). We found no statistically significant difference in transmissibility between symptomatic and asymptomatic individuals. Further, household contacts were associated with a significantly larger risk of SARS-CoV-2 infection than other types of contact. The GLMM model suggests two other statistically significant risk factors: the generation in the transmission chain and the number of contacts identified for an infector (Tab. 2). In particular, the transmission risk in the first generation was significantly higher than the later generations, possibly due to improved case isolation and contacts quarantine that deplete the number of susceptible individuals in the cluster. The same results were observed when accounted for the time period of the epidemic (Supporting Information, Tab. S10 and S11). We also found a slight but significant decrease in transmission risk from cases who reported more contacts. The inclusion of other potential risk factors, such as the gender of infectors and the gender of the contacts were not statistically significant, did not modify the estimated odds ratios for the other variables, and did not improve the fit of the model (Tab. S9, Tab. S10, and Fig. S7).

## Discussion

This analysis of SARS-CoV-2 transmission patterns and risk factors in Hunan, China, is based on the largest contact tracing dataset considered thus far. We found no difference in transmissibility between symptomatic and asymptomatic individuals and between age groups, while susceptibility to SARS-CoV-2 infection increased with age. We provide evidence of both pre-symptomatic and asymptomatic SARS-CoV-2 transmission, with the former potentially accounting for up to 59.2% of all transmission events in this dataset. In addition, we estimate that SARS-CoV-2 transmission in households is responsible for most of secondary and tertiary infections. Further, within a cluster, individuals who were exposed to primary cases experienced a significantly higher risk of SARS-CoV-2 infection than those exposed to later cases.

The exposure history data used in this study were collected from in-depth epidemiological investigations, allowing us to provide robust estimation of several key time-to-event distributions. Previous estimates suffered of large uncertainty, ranging from 3.0 days to 7.8 days for the serial interval ^1,17-22^ and from 4.8 days to 8.0 days for the incubation period ^1,23-28^. We note that short estimates of the serial interval such as the one obtained for Brazil ^21^ tend to be skewed as secondary cases tend to recall more recent contacts, which is especially true when a major epidemic is unfolding ^29^. This appears not to be the case in Hunan where the exponential growth phase of the outbreak lasted only about two weeks ^23^ and the effort heavily relied on forward contact tracing. Still, our estimates fall within these intervals. Moreover, in agreement with Sheikh et al ^30^, we found that the mean serial interval shortened over time, reflecting increased timeliness of case isolation that truncates successful onward transmission. Unlike the serial interval and the incubation period, only a few studies ^31,32^ provide estimates of the generation time, as it is hard to directly infer from field investigations given that it requires information on the infection dates of both the infector and her/his infectees. In this work, we estimate the mean generation time at 5.7 days (median: 5.5, SD: 1.8), in general agreement with Ferretti, et al ^32^(median: 5.0 days; SD: 1.9 days). Solid estimates of the generation time are key as, in conjunction with epidemic growth rate, they can be used to estimate the reproduction number of an epidemic ^33,34^. In the absence of such data, many studies so far have relied on the distribution of the serial interval as an approximation of the generation time ^1,35^. However, individual variability in the duration of the incubation period is expected to widen the distribution of the serial interval with respect to that of the generation time. This is highlighted by the IQR of the two distributions estimated here, namely the mean of the serial interval was estimated at 5.5 days (IQR: 0.9-9.4) and that of the generation time at 5.7 days (IQR: 4.5-6.8).

Previous studies show a relatively high proportion of pre-symptomatic transmission, but estimates vary significantly, ranging between 13-62% ^1,32,36^. Our estimate (59.2%) nears the high end of the range found in the literature. This may be due to two main factors. First, the fraction of pre-symptomatic transmission heavily depends on the intensity of contact tracing and isolation strategy (e.g., whether cases are promptly isolated in dedicated facilities at the time of symptom onset or are isolated at home). Second, the depth of the contact tracing investigation may determine the rate of ascertainment of index cases. Our analysis suggests a key role of interventions (e.g., contact tracing and case isolation) in decreasing the risk of infection, as the risk of infection decreased with the number of the generations in the transmission chain. As discussed in previous studies ^15,30,31^, the effectiveness of tracing and isolation/quarantine heavily depends on a quick identification of cases. Here we estimated the mean time interval from symptom onset to PCR sample collection and to laboratory confirmation to be 4.7 and 6.4 days, respectively. However, contacts were quarantined preventively before the diagnosis was confirmed. Finally, it is important to note that, by definition, our analysis includes only contacts occurring up to 2 days prior symptom onset of the presumed infector (as per Chinese authorities’ policy ^37^). This may potentially lead to underestimation of pre-symptomatic transmission outside the household and skew the distribution of infectiousness. Future analyses of viral load data may provide further support to our estimates of the infectiousness profile over time.

We found evidence of asymptomatic transmission in several clusters, with 15 secondary cases (out of 432 transmission events) linked to asymptomatic infectors, similar to Chen et al (6/132 events) ^38^ and Liu et al (24/914 events) ^39^. Other studies provide evidence of asymptomatic infection ^12,36,40^, but do not attempt to quantify its contribution to transmission. Our multivariate analysis shows no statistically significant difference in the transmissibility between symptomatic and asymptomatic individuals. This highlights that the low proportion of cases generated by asymptomatic individuals in this study (3.5%) can be partially explained by the lower probability of identification of asymptomatic index cases. In fact, by considering only clusters with at least one asymptomatic transmission event, the proportion of asymptomatic transmission increases to 60% (15/25). However, it is important to note our data cannot be used to estimate the probability of developing symptoms as a fraction of asymptomatic infections (e.g., entire clusters that consists only of asymptomatic subjects) may have been missed despite extensive PCR testing performed by the Hunan CDC. In fact, testing focused on symptomatic contacts before February 7, 2020, and was expanded to all contacts afterwards. Therefore, our findings cannot be used to quantitatively estimate the percentage of infected individuals who develop symptoms.

In agreement with previous studies, we found that the risk of infection from a household member is larger than that resulting from other contacts ^10,41^. This may be explained by the duration, type, and frequency of contacts between household members as well as the impact of interventions (such as household quarantine) on household contacts. Consistent with the transmissibility of H1N1pdm influenza during the 2009 pandemic in the US ^42^, we found that SARS-CoV-2 transmissibility decreased with the number of contacts, although the effect is small. Further cohort studies are needed to explain this connection, possibly recording number, type, and duration of contacts. It is important to stress that the observed significantly higher risk of infection in households calls for measures targeted at households, such as providing isolation shelters for mild cases that can remove SARS-CoV-2 infectors from households and thereby interrupt chains of within-household transmission ^43,44^. Although we estimated a higher susceptibility to SARS-CoV-2 infection among the elderly, this finding has to be cautiously interpreted. This findings may stem from a higher probability of infection detection among the elderly due to higher probability of developing symptoms and present severe illness ^45^.

Despite the challenges of reporting a low number of infections among children and the complexity of establishing epidemiologic links between children and adults within households ^19^, we assessed the effects of infector and infectee demographics and other characteristics on SARS-CoV-2 susceptibility and infectivity. We found that the odds of infection was significantly higher for first-generation infectors than for later generation ones. Together with a small number of infectious children in the first generation, this contributed to a lower total number of infections generated by children (see Tab. S11). However, when accounting for all confounding factors, including generation number, we found no statistical evidence of differential transmissibility by age group (Tab. 2). Interestingly, while younger individuals typically have more contacts than other age groups both in China ^9,46,47^ (range: 18.2-22.3 contacts per day) and elsewhere ^9,48-52^, the number of individual contacts reported by each infectious child in contact tracing data was considerably lower (mean: 7.7) during the outbreak in Hunan. Such a marked reduction in contacts was likely connected with the interventions in place (lockdown policy) and school closures (either for the New Year vacation and later as part of interventions). Therefore, caution should be applied when evaluating policies that increase the number of contacts among children, such as re-opening schools or summer camps. In addition, our findings suggest that the risk of acquiring SARS-CoV-2 infection steadily increases with age (in agreement with Zhang, et al.^9,11^). Nonetheless, it is important to remark that our estimates of the infectiousness by age groups are based on a small sample size of younger individuals. Further studies are needed to confirm our finding.

Our study is not without limitations. First, it suffers from the classic limitations of any epidemiological field investigation. Despite the longitudinal and in-depth investigation of each case and her/his contacts, we could not always accurately reconstruct the entire transmission chain and fully avoid recall bias in individual records. Also, the imperfect sensitivity of PCR testing should be taken into consideration, especially as it highly depends on the delay between the time of infection and specimen collection ^53^. Unfortunately, we do not have a representative sample for the date of sample collection, thus we cannot correct for this factor, possibly leading to an underestimation of the number of SARS-CoV-2 infected asymptomatic individuals. Moreover, we cannot rule out the possibility of indirect exposures (e.g., contaminated surfaces), which may affect the identification of epidemiological links. High-resolution genomic and virologic surveillance data would be needed to decrease the uncertainty on the links identified by the epidemiological investigation and to better distinguish direct vs. community transmission ^54,55^. Second, the duration of per-contact exposure was not reported in the dataset and we were thus unable to correct for this factor. This may contribute to explain the importance attributed to household contacts in our regression analysis and why individuals with more contacts have lower transmission risk per contact. Third, despite controlling known factors associated with transmissibility, we cannot exclude the possibility that there are other potential factors that may confound the estimated effect of current covariates.

In conclusion, the evidence of pre-symptomatic and asymptomatic SARS-CoV-2 transmission shown in this study underlines the key role of undetectable SARS-CoV-2 transmission that can hinder control efforts. Control measures should thus be tailored accordingly, especially contact tracing, testing, and isolation. Our findings show a high risk of acquiring SARS-CoV-2 from infected individuals not showing symptoms (either because pre-symptomatic or asymptomatic), thus supporting the enhancement of personal precautions such as wearing a mask and improved hygiene practice. In addition, school reopening, and the consequent increase in the number of daily contacts among children and teenagers, is expected to increase the contribution of children to SARS-CoV-2 transmission. School outbreaks have already been reported in several occasions ^5,56-58^; time will tell whether schools become a major foci of transmission.

**Table 1.**
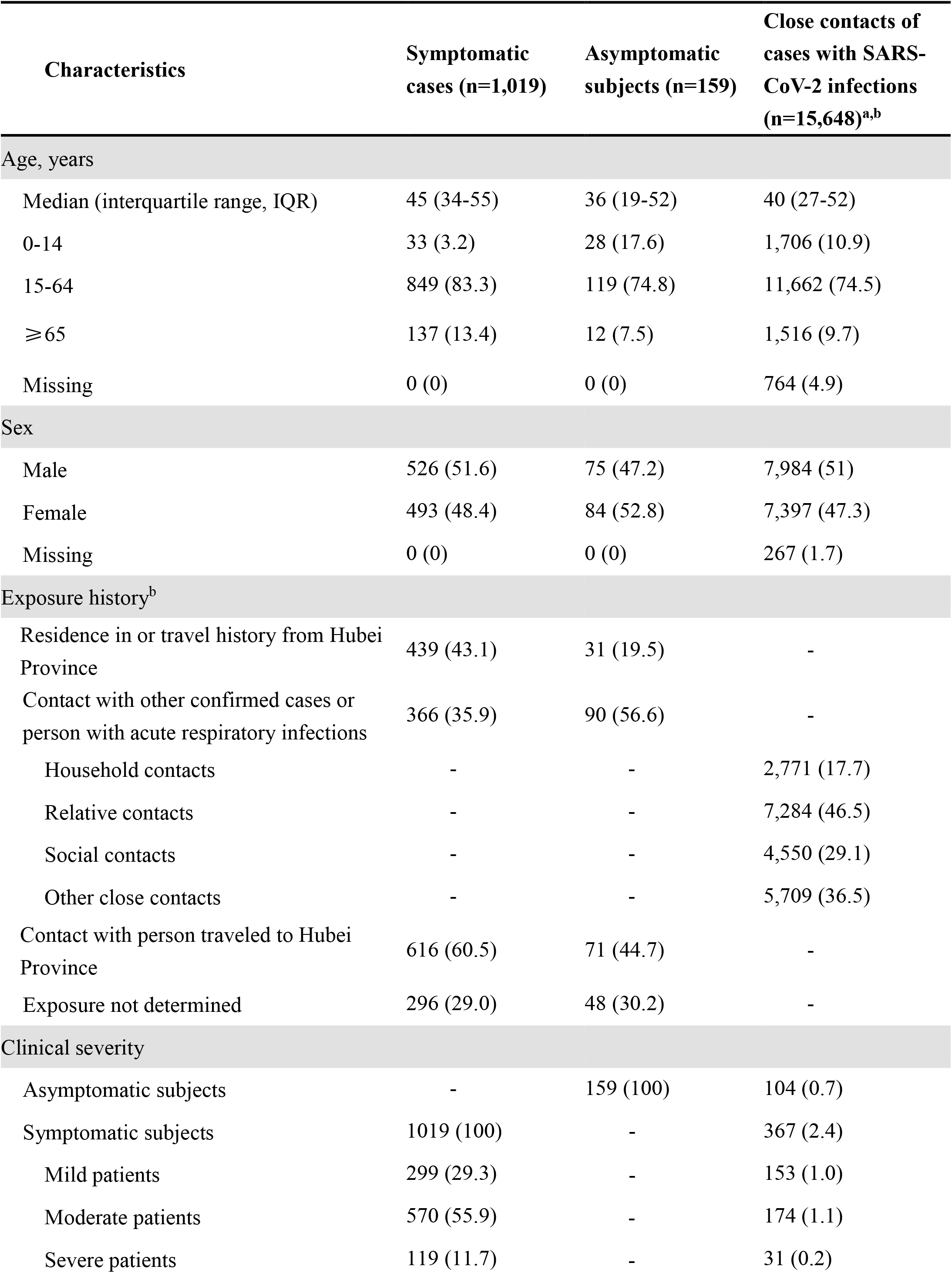
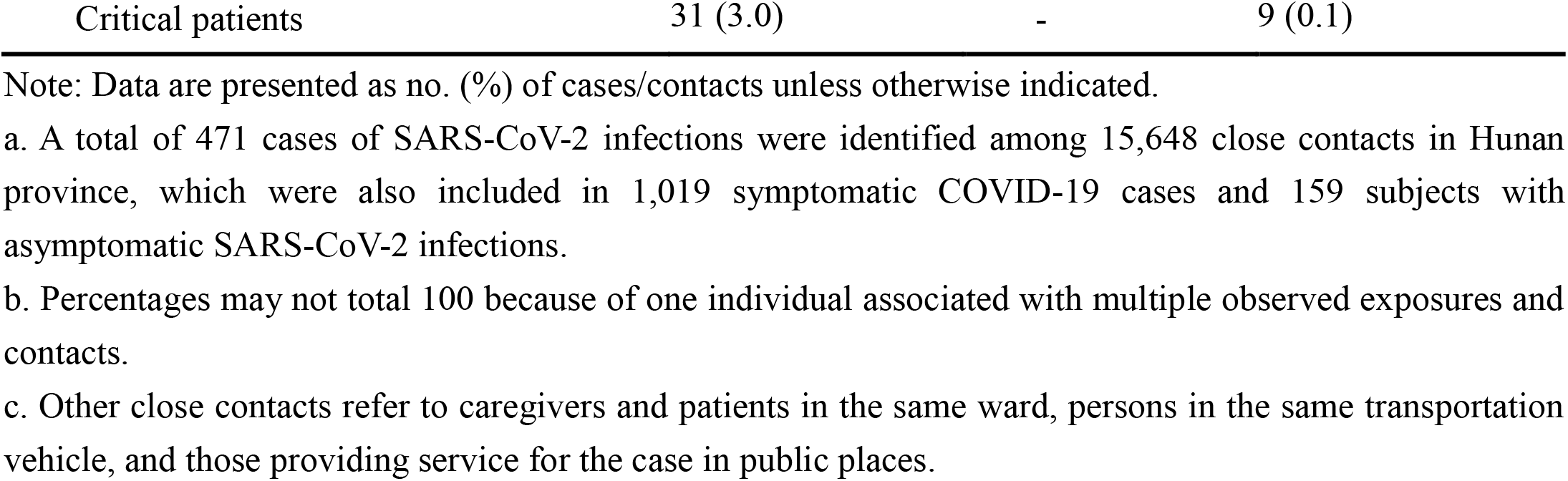
Characteristics of symptomatic cases, asymptomatic subjects, and their close contacts in Hunan Province, China

| Characteristics | Symptomatic cases (n=1,019) | Asymptomatic subjects (n=159) | Close contacts of cases with SARS-CoV-2 infections (n=15,648)<sup>a,b</sup> |
| --- | --- | --- | --- |
| Age, years |  |  |  |
| Median (interquartile range, IQR) | 45 (34-55) | 36 (19-52) | 40 (27-52) |
| 0-14 | 33 (3.2) | 28 (17.6) | 1,706 (10.9) |
| 15-64 | 849 (83.3) | 119 (74.8) | 11,662 (74.5) |
| ≥65 | 137 (13.4) | 12 (7.5) | 1,516 (9.7) |
| Missing | 0 (0) | 0 (0) | 764 (4.9) |
| Sex |  |  |  |
| Male | 526 (51.6) | 75 (47.2) | 7,984 (51) |
| Female | 493 (48.4) | 84 (52.8) | 7,397 (47.3) |
| Missing | 0 (0) | 0 (0) | 267 (1.7) |
| Exposure history<sup>b</sup> |  |  |  |
| Residence in or travel history from Hubei Province | 439 (43.1) | 31 (19.5) | - |
| Contact with other confirmed cases or person with acute respiratory infections | 366 (35.9) | 90 (56.6) | - |
| Household contacts | - | - | 2,771 (17.7) |
| Relative contacts | - | - | 7,284 (46.5) |
| Social contacts | - | - | 4,550 (29.1) |
| Other close contacts | - | - | 5,709 (36.5) |
| Contact with person traveled to Hubei Province | 616 (60.5) | 71 (44.7) | - |
| Exposure not determined | 296 (29.0) | 48 (30.2) | - |
| Clinical severity |  |  |  |
| Asymptomatic subjects | - | 159 (100) | 104 (0.7) |
| Symptomatic subjects | 1019 (100) | - | 367 (2.4) |
| Mild patients | 299 (29.3) | - | 153 (1.0) |
| Moderate patients | 570 (55.9) | - | 174 (1.1) |
| Severe patients | 119 (11.7) | - | 31 (0.2) |

Note: Data are presented as no. (%) of cases/contacts unless otherwise indicated.
a. A total of 471 cases of SARS-CoV-2 infections were identified among 15,648 close contacts in Hunan province, which were also included in 1,019 symptomatic COVID-19 cases and 159 subjects with asymptomatic SARS-CoV-2 infections.
b. Percentages may not total 100 because of one individual associated with multiple observed exposures and contacts.
c. Other close contacts refer to caregivers and patients in the same ward, persons in the same transportation vehicle, and those providing service for the case in public places.

**Table 2.**
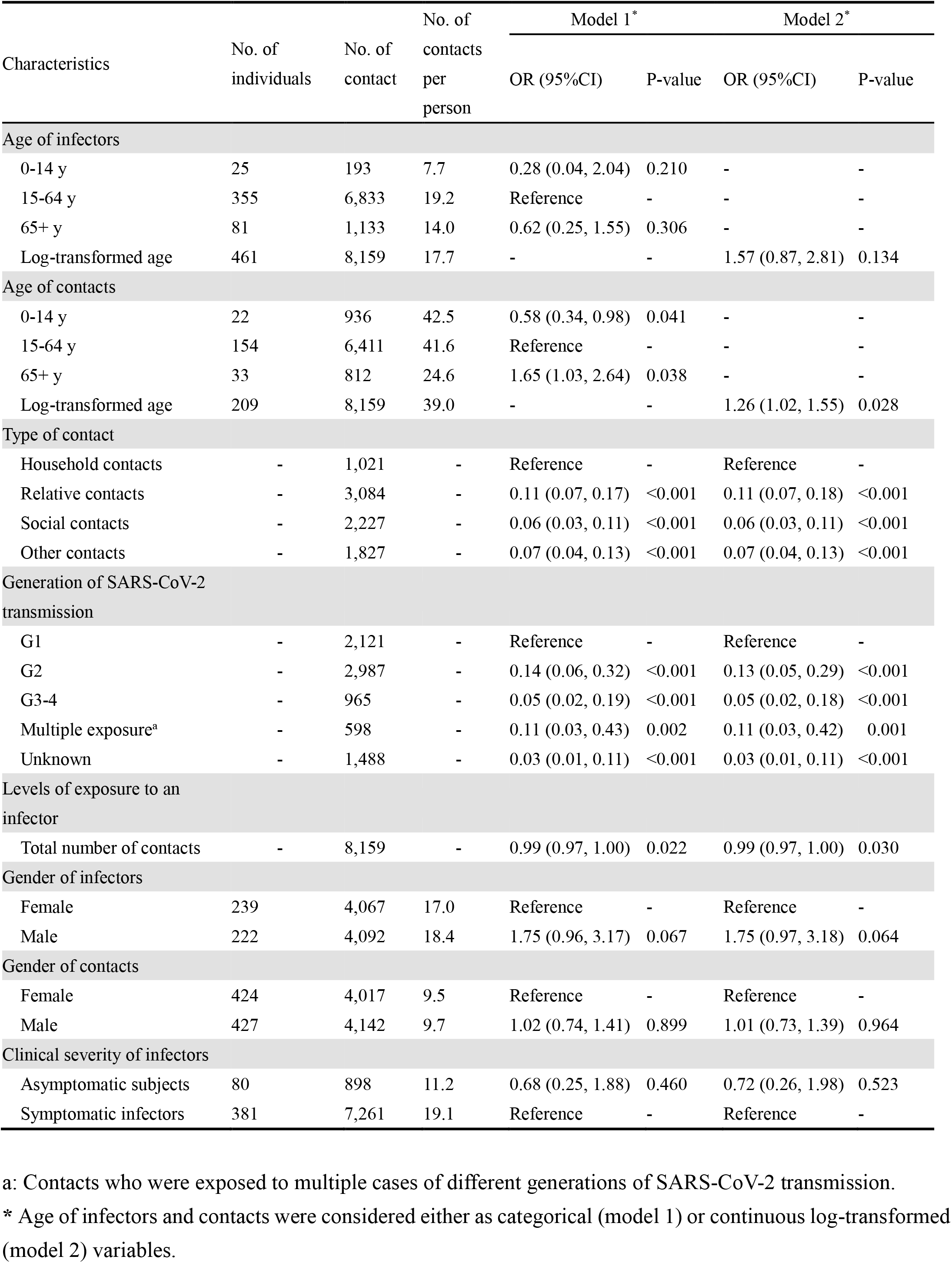
Estimating the association of demographic and behavioral factors with the risk of acquiring and transmitting SARS-CoV-2

## Methods

### COVID-19 surveillance system, field epidemiological investigations, and contact tracing

In response to the COVID-19 outbreak, in late December 2019, the Chinese Center for Disease Control and Prevention (China CDC) launched a new surveillance system for COVID-19 cases. A description of the surveillance system is reported elsewhere ^1^. On January 21, 2020, the first COVID-19 case was confirmed in Hunan Province. Since then, active field epidemiological investigations of suspected or confirmed SARS-CoV-2 infections as well as their contacts have been initiated.

The definition of suspected and confirmed COVID-19 cases (i.e., symptomatic individuals), as well as subjects with asymptomatic SARS-CoV-2 infections (i.e., asymptomatic subjects) was based on the New Coronavirus Pneumonia Prevention and Control Program published by the National Health Commission (NHC) of China and the World Health Organization (WHO) ^59^. A suspected COVID-19 case was defined as a person who met one or more clinical criteria and had an epidemiological link to SARS-CoV-2 positive individuals or history of travel to/from regions reporting widespread SARS-CoV-2 transmission (Supporting Information, p2). A confirmed COVID-19 case was defined as a suspected case with positive real-time RT-PCR results, while an asymptomatic subject was defined as an individual with laboratory confirmation of SARS-CoV-2 infection, but without any clinical symptom (e.g., no fever or cough) within the quarantine/observation period (i.e., 14 days). Confirmed COVID-19 cases were categorized by clinical severity, including mild, moderate, severe and critical illnesses (as defined in Supporting Information, Tab. S1).

SARS-CoV-2 infected individuals were identified using a variety of measures. In particular, i) to identify travel-associated cases, traffic entrance and community screening were performed in high-risk populations who had a history of traveling from/to Wuhan City/Hubei Province; ii) to identify symptomatic cases, passive surveillance in hospitals and outpatient practices were monitored; iii) to capture potential symptomatic cases and asymptomatic subjects, systematic tracing and monitoring of contacts of confirmed cases was performed. In particular, screening measures i) and iii) were used to identify SARS-CoV-2 infected asymptomatic individuals. Once a suspected or confirmed COVID-19 case was identified, a field epidemiology investigation was undertaken by the local CDC. Data were collected on demographic characteristics, clinical symptoms, and activity patterns starting 14 days before symptom onset and until confirmation or isolation in the hospital. All cases detected between January 16 and April 02, 2020 were interviewed using a standardized questionnaire. In addition, each individual with suspected or confirmed SARS-CoV-2 infection was asked to provide a list of locations she/he visited (e.g., workplace, health-care facilities) and her/his contacts. On the basis of this list, active contact tracing was then initiated by the investigation team. Screening interviews, checking of travel records based on public security cameras and traffic system, and digital health records were also collected to assess whether an individual met the definition of close contact. Once a close contact was identified and traced, she/he was quarantined at a designated place (e.g., hotel room) or at home and followed up for 14 days ^59^. Close contacts were interviewed using a standardized form before they were quarantined. The form comprised basic demographic information (e.g., age and sex), and detailed a record of the timing, frequency, and type of exposures to the case(s) who triggered the investigation. An earlier version of the data from contact tracing operations containing only reduced descriptive information on contacts was used for the estimation of age-specific susceptibility in Zhang, et al.^9^.

### Specimen collection and laboratory testing

Upper respiratory specimens (nasopharyngeal and oropharyngeal swabs) were collected from all suspected cases as well as their close contacts. Before February 7, 2020 specimens were collected for testing from each close contact if she/he developed symptoms during quarantine period. After February 7, 2020, specimens were collected at least once during quarantine, regardless of symptoms. After January 27, the designated hospitals and local CDCs were approved to conduct real-time RT-PCR assay for diagnosis of COVID-19 using a standardized laboratory testing procedure according to the “Novel coronavirus pneumonia Diagnosis and Treatment Program” released by NHC of China. The assays were performed in laboratory equipped with BSL-2 facilities (Supporting Information, p3-4).

### Close contacts, sporadic cases, and clusters

Close contacts were defined as individuals who had close-proximity interactions (within 1 meter) with clinically suspected and laboratory-confirmed SARS-CoV-2 cases, for the period from 2 days before, to 14 days after, the potential infector’s symptom onset. For those exposed to asymptomatic subjects, the contact period was from 2 days before, to 14 days after, a respiratory sample was taken for real-time RT-PCR testing. Close contacts included, but were not limited to, household contacts (i.e., household members regularly living with the case), relatives (i.e., family members who had close contacts with the case but did not live with the case), social contacts (i.e., a work colleague or classmate), and other close contacts (i.e., caregivers and patients in the same ward, persons sharing a vehicle, and those providing a service in public places, such as restaurants or movie theatres) ^37^.

A cluster of SARS-CoV-2 infections was defined as a group of two or more confirmed cases or asymptomatic subjects with an epidemiologic link (Supporting Information, p3). Epidemiologically linked cases were classified according to the generation time of SARS-CoV-2 transmission and the setting where exposure took place, with primary cases considered as first generation. A sporadic case was defined as a confirmed case of SARS-CoV-2 infection (either symptomatic or asymptomatic) who did not belong to any of the reported clusters.

We define pre-symptomatic transmission as a direct transmission event that takes place before the date of symptom onset of the infector, while asymptomatic transmission is a transmission event from a person who did not develop symptoms within the quarantine/observation period.

## Data Availability

Individual-based data on 1,178 SARS-CoV-2 infected individuals and their 15,648 contacts identified by contact tracing monitoring over the period from January 13-April 02, 2020 were extracted from the notifiable infectious diseases reporting system in Hunan Province, China. Demographic characteristics, severity classification, exposure and travel history, and key clinical timelines were retrieved.

## Data Availability

All relevant data are within the manuscript and its Supporting Information files.

## Statistical analysis

We provide descriptive statistics of the characteristics of cases and their close contacts, including demographic factors and exposures (Supporting Information, p5-p7). We estimated the incubation period (i.e., the time delay from infection to illness onset), the serial interval (i.e., the time interval between the onset of symptoms in a primary case and in her/his secondary cases), the generation time (i.e., the time interval between infection of the primary case and of her/his secondary cases), and the infectiousness profile (i.e., the daily distribution of the probability of transmission since the date of symptom onset; see ^1,60^ and Supporting Information, p8-p12 for methods). We also estimated the interval from symptom onset to the sampling date of first PCR and to laboratory confirmation by using a maximum likelihood estimator and fitting three distributions (Weibull, gamma, and lognormal) (Supporting Information, p12). The goodness of fit was assessed using Akaike information criterion (AIC). We restrict the estimation of incubation period to 268 locally acquired infections with information on both the date(s) of exposure and generation of SARS-CoV-2 transmission in the cluster.

We rely on the contact tracing data to describe the age-specific contact matrices for SARS-CoV-2 infectors and their contacts (Supporting Information, p11), and we present the number of contacts per person by demographical characteristics of SARS-CoV-2 infectors and their contacts. To focus on local transmission pairs with clearly epidemiological links, we excluded those travel-related cases and their successive cases (as people in the same cluster often share the same travel history, making hard to disentangle the transmission chain). Additionally, generalized linear mixed-effects model, GLMM, for binary data with logit link were built to quantify the effects of potential drivers of susceptibility and infectivity of the SARS-CoV-2 virus (i.e., odds ratio and marginal effect), based on 8,159 individual records of contacts who were exposed to locally transmitted cases (see Supporting Information, p11-12). These risk factors include age and gender of infectors/contacts, type of contact, generation of SARS-CoV-2 transmission in a cluster, as well as the number of contacts of an infector. Statistical analyses were performed using the R software, version 3.5.0.

## Ethical approval statement

This study was approved by the ethic committee of the Hunan CDC with a waiver of informed consent due to a public health outbreak investigation (IRB No. 2020005).

## Data availability

All aggregated data analyzed in this study are included in the Article and Supplementary Information. All data supporting the findings of this study are available from the authors upon request.

## Acknowledgments

National Science Fund for Distinguished Young Scholars (No. 81525023), National Science and Technology Major Project of China (No. 2017ZX10103009-005, No. 2018ZX10713001-007, No. 2018ZX10201001-010), and Hunan Provincial Innovative Construction Special Fund: Emergency response to COVID-19 outbreak (No. 2020SK3012).

## Author contributions

S. Hu, W. Wang, Y. Wang, L. Gao, and H. Yu had full access to all of the data in the study and take responsibility for the integrity of the data and the accuracy of the data analysis. L. Gao, M. Ajelli, and H. Yu were responsible for its conception and design. K. Luo, L. Ren, Q. Sun, X. Chen, G. Zeng, J. Li, L. Liang, Z. Deng, W. Zheng, M. Li, H. Yang, J. Guo, K. Wang, X. Chen, Z. Liu, H. Yan, H. Shi, Z. Chen, and Y. Zhou were responsible for the acquisition, analysis, or interpretation of data. S. Hu, W. Wang, M. Litvinova, M. Ajelli and H. Yu drafted the manuscript. K. Sun, A. Vespignani, C. Viboud, L. Gao, M. Ajelli, H. Yu made critical revision of the manuscript for important intellectual content. W. Wang, Y. Wang, M. Litvinova, and M. Ajelli did the data analysis. K. Luo, Q. Sun, G. Zeng, Z. Deng, H. Yang, Z. Liu, and K. Sun provided administrative, technical, or material support.

## Additional information

**Supplementary Information** is available for this paper. Requests for materials should be addressed to L.G. and H.Y.

## Reference

1. Li, Q., et al. Early Transmission Dynamics in Wuhan, China, of Novel Coronavirus–Infected Pneumonia. New England Journal of Medicine (2020).

2. World Health Organization. Coronavirus disease (COVID-19) pandemic. (2020).

3. World Health Organization. Coronavirus disease 2019 (COVID-19) Situation Report – 170. Vol. 2020 (2020).

4. Van Lancker, W. & Parolin, Z. COVID-19, school closures, and child poverty: a social crisis in the making. The Lancet Public Health 5, e243–e244 (2020).

5. United Nations Educational, S.a.C.O. Education: From disruption to recovery. Vol. 2020 (2020).

6. Sinha, I.P., et al. COVID-19 infection in children. The Lancet Respiratory Medicine 8, 446–447 (2020).

7. Poletti, P., et al. Probability of symptoms and critical disease after SARS-CoV-2 infection. in arXiv e-prints 2006.08471 (2020).

8. Pollán, M., et al. Prevalence of SARS-CoV-2 in Spain (ENE-COVID): a nationwide, population-based seroepidemiological study. The Lancet.

9. Zhang, J., et al. Changes in contact patterns shape the dynamics of the COVID-19 outbreak in China. Science 368, 1481–1486 (2020).

10. Jing, Q.L., et al. Household secondary attack rate of COVID-19 and associated determinants in Guangzhou, China: a retrospective cohort study. Lancet Infect Dis (2020).

11. Wu, J.T., et al. Estimating clinical severity of COVID-19 from the transmission dynamics in Wuhan, China. Nat Med 26, 506–510 (2020).

12. Oran, D.P. & Topol, E.J. Prevalence of Asymptomatic SARS-CoV-2 Infection: A Narrative Review. Ann Intern Med (2020).

13. Baggett, T.P., Keyes, H., Sporn, N. & Gaeta, J.M. Prevalence of SARS-CoV-2 Infection in Residents of a Large Homeless Shelter in Boston. JAMA (2020).

14. Arons, M.M., et al. Presymptomatic SARS-CoV-2 Infections and Transmission in a Skilled Nursing Facility. N Engl J Med 382, 2081–2090 (2020).

15. Cheng, H.-Y., et al. High transmissibility of COVID-19 near symptom onset. medRxiv, 2020.2003.2018.20034561 (2020).

16. Cereda, D., et al. The early phase of the COVID-19 outbreak in Lombardy, Italy. in arXiv e-prints 2003.09320 (2020).

17. Nishiura, H., Linton, N.M. & Akhmetzhanov, A.R. Serial interval of novel coronavirus (COVID-19) infections. Int J Infect Dis 93, 284–286 (2020).

18. Du, Z., et al. Serial Interval of COVID-19 among Publicly Reported Confirmed Cases. Emerging Infectious Disease journal 26, 1341 (2020).

19. Danis, K., et al. Cluster of coronavirus disease 2019 (Covid-19) in the French Alps, 2020. Clinical Infectious Diseases (2020).

20. Ali, S.T., et al. Serial interval of SARS-CoV-2 was shortened over time by nonpharmaceutical interventions. Science (2020).

21. Prete, C.A., Jr., et al. Serial interval distribution of SARS-CoV-2 infection in Brazil. Journal of Travel Medicine (2020).

22. You, C., et al. Estimation of the time-varying reproduction number of COVID-19 outbreak in China. International Journal of Hygiene and Environmental Health 228, 113555 (2020).

23. Zhang, J., et al. Evolving epidemiology and transmission dynamics of coronavirus disease 2019 outside Hubei province, China: a descriptive and modelling study. Lancet Infect Dis 20, 793–802 (2020).

24. Linton, N.M., et al. Incubation Period and Other Epidemiological Characteristics of 2019 Novel Coronavirus Infections with Right Truncation: A Statistical Analysis of Publicly Available Case Data. J Clin Med 9(2020).

25. Backer, J.A., Klinkenberg, D. & Wallinga, J. Incubation period of 2019 novel coronavirus (2019-nCoV) infections among travellers from Wuhan, China, 20-28 January 2020. Euro Surveill 25(2020).

26. Bi, Q., et al. Epidemiology and transmission of COVID-19 in 391 cases and 1286 of their close contacts in Shenzhen, China: a retrospective cohort study. The Lancet Infectious Diseases (2020).

27. Tian, S., et al. Characteristics of COVID-19 infection in Beijing. J Infect 80, 401–406 (2020).

28. Xu, T., et al. Clinical features and dynamics of viral load in imported and non-imported patients with COVID-19. Int J Infect Dis 94, 68–71 (2020).

29. Park, S.W., Champredon, D. & Dushoff, J. Inferring generation-interval distributions from contact-tracing data. J R Soc Interface 17, 20190719 (2020).

30. Ali, S.T., et al. Serial interval of SARS-CoV-2 was shortened over time by nonpharmaceutical interventions. Science 369, 1106 (2020).

31. Ganyani, T., et al. Estimating the generation interval for coronavirus disease (COVID-19) based on symptom onset data, March 2020. Euro Surveill 25(2020).

32. Ferretti, L., et al. Quantifying SARS-CoV-2 transmission suggests epidemic control with digital contact tracing. Science 368(2020).

33. Wallinga, J. & Lipsitch, M. How generation intervals shape the relationship between growth rates and reproductive numbers. Proc Biol Sci 274, 599–604 (2007).

34. Cori, A., Ferguson, N.M., Fraser, C. & Cauchemez, S. A New Framework and Software to Estimate Time-Varying Reproduction Numbers During Epidemics. American Journal of Epidemiology 178, 1505–1512 (2013).

35. Zhao, S., et al. Serial interval in determining the estimation of reproduction number of the novel coronavirus disease (COVID-19) during the early outbreak. Journal of Travel Medicine 27(2020).

36. Kimball, A., et al. Asymptomatic and Presymptomatic SARS-CoV-2 Infections in Residents of a Long-Term Care Skilled Nursing Facility - King County, Washington, March 2020. MMWR Morb Mortal Wkly Rep 69, 377–381 (2020).

37. National Health Commission of the People’s Republic of China. The prevention and control of 2019 Novel Coronavirus Pneumonia (7th Edition). Vol. 2020 (2020).

38. Chen Yi, W.A., Yi Bo, Ding Keqin, Wang Haibo, Wang Jianmei, Shi Hongbo, Wang Sijia, Xu Guozhang. The epidemiological characteristics of infection in close contacts of COVID-19 in Ningbo city. Chinese Journal of Epidemiology 41(2020).

39. Liu, Z., Chu, R., Gong, L., Su, B. & Wu, J. The assessment of transmission efficiency and latent infection period on asymptomatic carriers of SARS-CoV-2 infection. Int J Infect Dis (2020).

40. Rothe, C., et al. Transmission of 2019-nCoV Infection from an Asymptomatic Contact in Germany. New England Journal of Medicine (2020).

41. Li, W., et al. The characteristics of household transmission of COVID-19. Clin Infect Dis (2020).

42. Cauchemez, S., et al. Household transmission of 2009 pandemic influenza A (H1N1) virus in the United States. N Engl J Med 361, 2619–2627 (2009).

43. Chen, S., et al. Fangcang shelter hospitals: a novel concept for responding to public health emergencies. The Lancet 395, 1305–1314 (2020).

44. Dickens, B.L., Koo, J.R., Wilder-Smith, A. & Cook, A.R. Institutional, not home-based, isolation could contain the COVID-19 outbreak. The Lancet 395, 1541–1542 (2020).

45. Poletti, P., et al. Probability of symptoms and critical disease after SARS-CoV-2 infection. 2006.08471 (2020).

46. Zhang, J., et al. Patterns of human social contact and contact with animals in Shanghai, China. Scientific Reports 9, 15141 (2019).

47. Guan, W.-j., et al. Clinical characteristics of 2019 novel coronavirus infection in China. medRxiv, 2020.2002.2006.20020974 (2020).

48. Johnstone-Robertson, S.P., et al. Social Mixing Patterns Within a South African Township Community: Implications for Respiratory Disease Transmission and Control. American Journal of Epidemiology 174, 1246–1255 (2011).

49. Ibuka, Y., et al. Social contacts, vaccination decisions and influenza in Japan. J Epidemiol Community Health 70, 162–167 (2016).

50. Kiti, M.C., et al. Quantifying Age-Related Rates of Social Contact Using Diaries in a Rural Coastal Population of Kenya. PLoS One 9, e104786 (2014).

51. Mossong, J., et al. Social contacts and mixing patterns relevant to the spread of infectious diseases. PLoS Med 5, e74 (2008).

52. Ajelli, M. & Litvinova, M. Estimating contact patterns relevant to the spread of infectious diseases in Russia. Journal of theoretical biology 419, 1–7 (2017).

53. Kucirka, L.M., Lauer, S.A., Laeyendecker, O., Boon, D. & Lessler, J. Variation in False-Negative Rate of Reverse Transcriptase Polymerase Chain Reaction-Based SARS-CoV-2 Tests by Time Since Exposure. Annals of internal medicine 173, 262–267 (2020).

54. Deng, X., et al. Case fatality risk of the first pandemic wave of novel coronavirus disease 2019 (COVID-19) in China. Clinical Infectious Diseases (2020).

55. Rockett, R.J., et al. Revealing COVID-19 transmission in Australia by SARS-CoV-2 genome sequencing and agent-based modeling. Nature Medicine 26, 1398–1404 (2020).

56. U.S. News. Hundreds of South Korean Schools to Close After Reopening. (2020).

57. 57. Yahoo News. Primary school forced to close and pupils and staff told to quarantine after catering team member gets COVID-19. (2020).

58. Stein-Zamir, C., et al. A large COVID-19 outbreak in a high school 10 days after schools’ reopening, Israel, May 2020. Euro Surveill 25, 2001352 (2020).

59. National Health Commission of the People’s Republic of China. Diagnosis and treatment guideline on pneumonia infection with 2019 novel coronavirus (6th trial edn). (2020).

60. Ashcroft, P., et al. COVID-19 infectivity pro1le correction. ArXiv e-prints (2020).

