## Supporting information for "Infectivity, susceptibility, and risk factors associated with SARS-CoV-2 transmission under intensive contact tracing in Hunan, China"

### Definitions of cases of SARS-CoV-2 infection

*Suspected COVID-19 cases*

A suspected COVID-19 case is defined as a person who meets three clinical criteria OR two clinical criteria and one of epidemiological criteria:

a) Clinical criteria: i) acute respiratory illness; ii) radiographic evidence of COVID-19 viral pneumonia; iii) normal or decreased white blood cells count in the early stage of the disease and normal or decreased lymphocyte count.

b) Epidemiological criteria: i) history of travel to or residence in Wuhan or domestic location reporting community transmission or countries/territories/areas/overseas reporting widespread SARS-CoV-2 transmission during the 14 days prior to symptom onset; ii) contact with any confirmed cases during the 14 days prior to symptom onset; iii) cluster of contact with COVID-19 patients (nucleic acids amplification test positive) within 14 days before symptom onset or to individuals with fever and/or symptoms of respiratory infection within 14 days.

*Clinical severity of COVID-19 confirmed cases*

We categorized confirmed COVID-19 cases according to their clinical severity, i.e., mild, moderate, severe, and critical case-patients. The details are presented in Tab. S1.

***Table S1.*** *Definitions of clinical severity of COVID-19 cases.* [1]

| **Clinical severity** | **Definition**[1] |
|---|---|
| Mild | Patients with mild symptoms, and no radiographic evidence of pneumonia |
| Moderate | Patients with fever, respiratory symptoms, and radiographic evidence of pneumonia |
| Severe | Patients had any of the following:<br>a. respiratory distress, breathing rate ≥30 beats/min; or<br>b. finger oxygen saturation ≤93% during resting state; or<br>c. $PaO_2/FiO_2$ ≤300mmHg (1mmHg = 0.133kPa). |

| | |
|---|---|
| | Patients whose pulmonary imaging have obvious progress of lesions (>50%) within 24~48 hours are managed by severe case. |
| Critical | Patients had any of the following:<br>a. respiratory failure and requires mechanical ventilation; or<br>b. shock; or<br>c. with other organ failures that requires ICU admission. |

*Epidemiologically-linked COVID-19 cases*

An individual with an epidemiologic link is a SARS-CoV-2 infected individual who has either been exposed to a symptomatic or an asymptomatic individual, or had the same exposure as the SARS-CoV-2 infected individuals. Generally, epidemiologically-linked cases include, but are not limited to SARS-CoV-2 infected individuals' household contacts (i.e., household members regularly living with the case), relatives (i.e., family members who had close contacts with the case but did not live with the case), social contacts (i.e., a work colleague or classmate), and other close contacts (i.e., caregivers and patients in the same ward, persons sharing a vehicle, and those providing a service for the case in public places) who have been close-proximity interactions (within 1 meter) with index case-patient and have acquired SARS-CoV-2 infections.

The flowchart describing the selection criteria of the analyzed subjects is shown in Fig. S1.

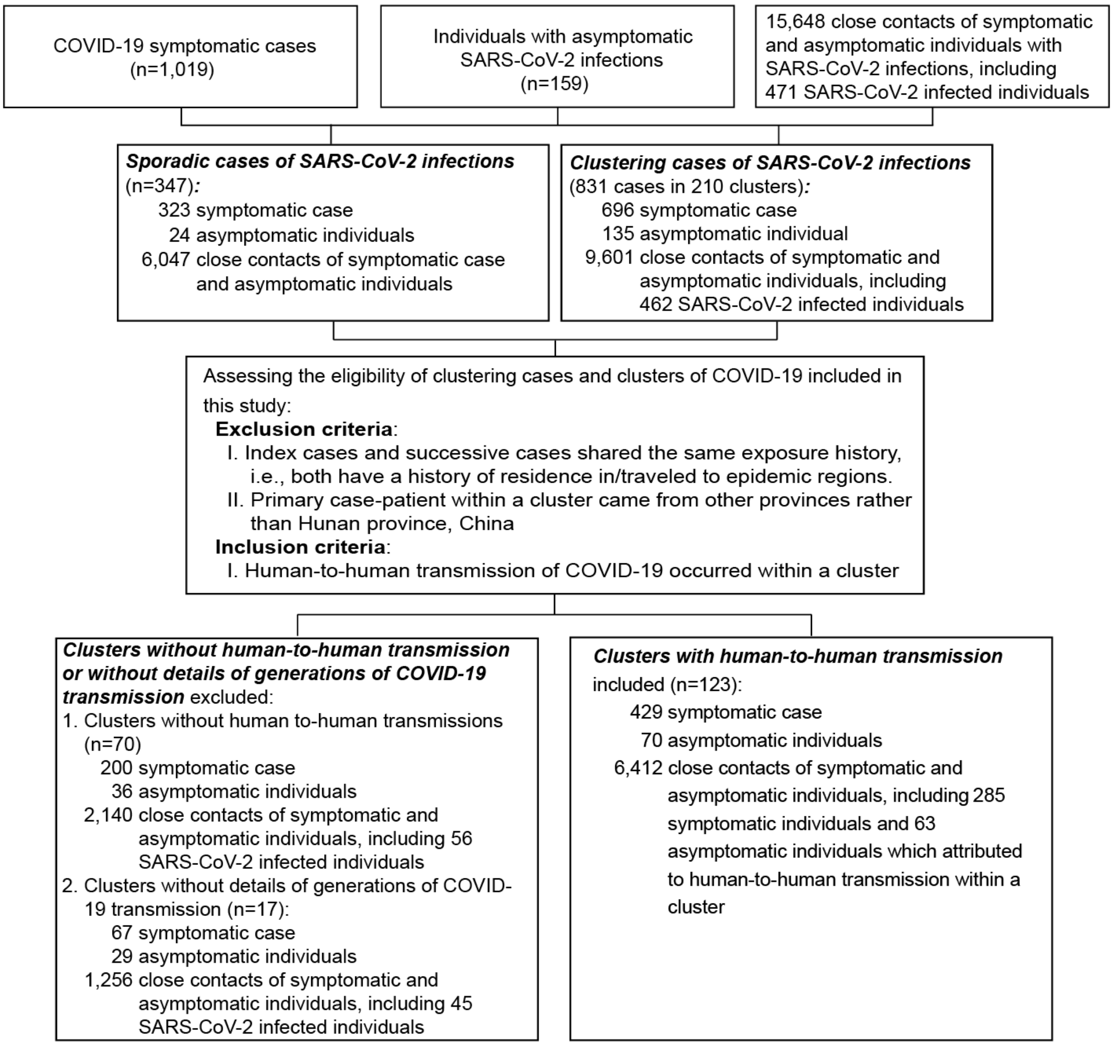

***Figure S1.*** *Flowchart describing the COVID-19 symptomatic cases, individual with asymptomatic infections, close contacts, and clusters with human-to-human transmission.*

### Specimen collection and laboratory testing

Since January 27, the designated hospitals and local Centers for Diseases Prevention and Controls (CDC) were approved to conduct real-time RT-PCR assay for diagnosis of COVID-19 using uniform laboratory testing procedures established by the World Health Organization (WHO). Total RNA was extracted using automated Nucleic Acid Extraction System 9600E (Xi'an TianLong Science and Technology Co., Ltd., Xi'an, China). Real-time reverse transcription polymerase chain reaction (RT-PCR) assay for SARS-CoV-2 was performed using a SARS-CoV-2 ORF1ab/N gene detection kit (Biogerm Medical Biotechnology Co.,Ltd, Shanghai, China), a product based on the recommendation of the National Institute for Viral Disease Control and Prevention, Chinese Center for Disease Control and Prevention. The open reading frame 1ab gene (ORF1ab) and nucleocapsid gene (N) were amplified and tested. Results were reported positive when both the ORF1ab gene and N gene were positive. Specimens tested as Ct-value of >=35 and <39.2 were retested for confirmation, a retest Ct-value of >=39.2 was treated as positive, otherwise negative.

### Overview of COVID-19 epidemics in Hunan Province, China

Overall, the dynamics of the epidemic in Hunan followed an exponential growth before January 23, 2020, and a decrease in the number of cases after February 1, 2020 (Fig. S2). Age descriptive statistics by other covariates are shown in Fig. S4.

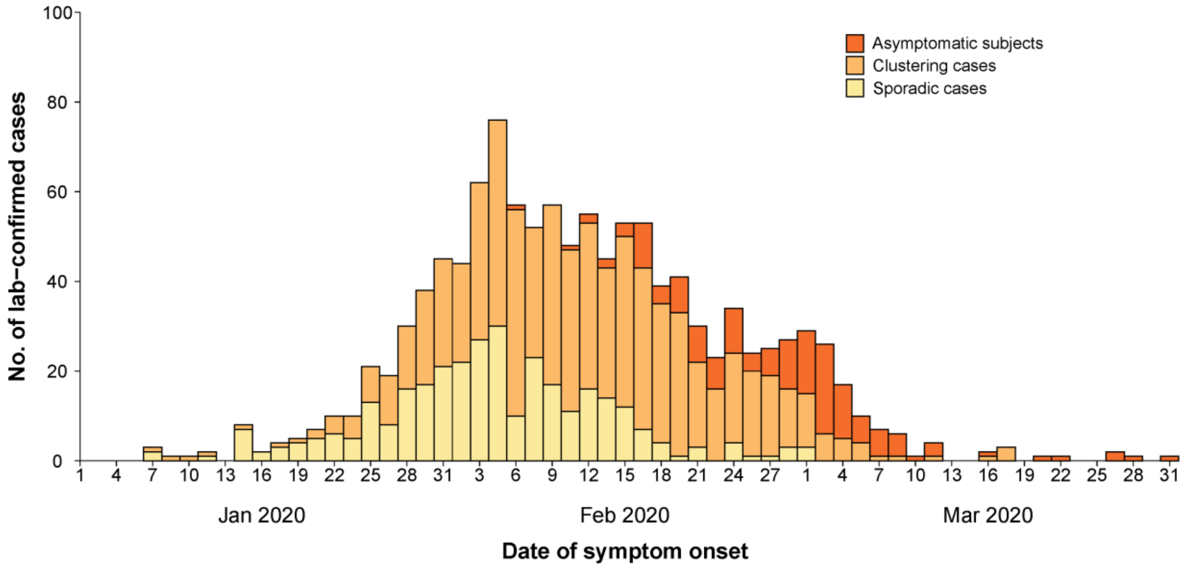

***Figure S2.*** *Epidemic curve of sporadic COVID-19 cases, individuals with asymptomatic SARS-COV-2 infections, and clustering COVID-19 cases in Hunan, China.*

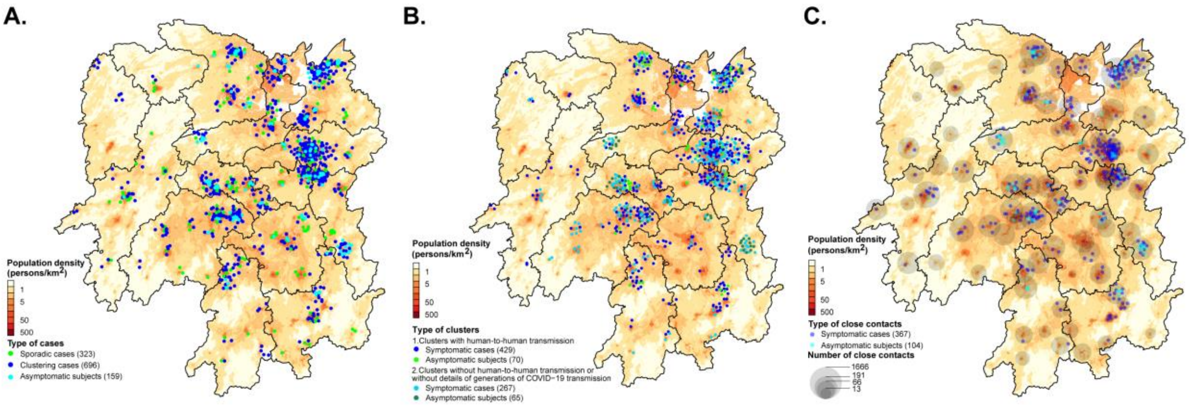

***Figure S3.*** *Geographical distribution of SARS-CoV-2 infected individuals and their close contacts stratified by the presence of symptoms and source of infection in Hunan Province, China. (A) Sporadic and clustered individuals with SARS-CoV-2 infections. (B) SARS-CoV-2 symptomatic and asymptomatic infected individuals by source of infection. (C) Close contacts of SARS-CoV-2 symptomatic and asymptomatic infected individuals.*

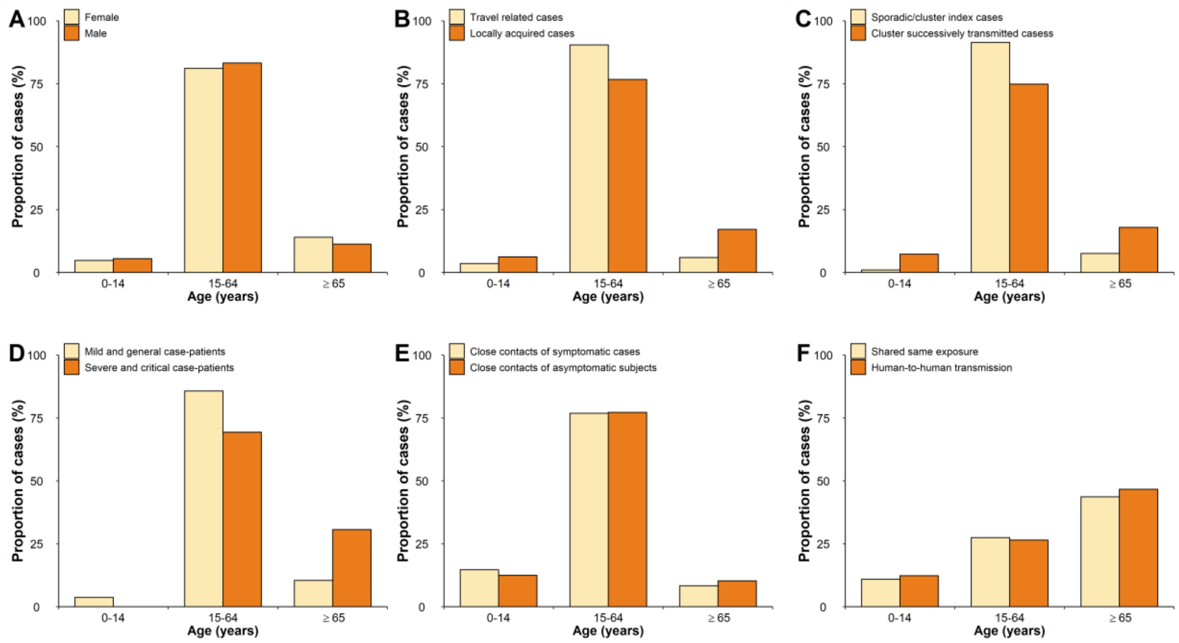

***Figure S4.*** *Age distribution of COVID-19 symptomatic cases, asymptomatic subjects as well as their close contacts. (A) Age and sex distribution of COVID-19 symptomatic cases. (B) Age distributions of travel related COVID-19 cases and locally acquired COVID-19 cases. (C) Age distributions of sporadic cases, index cases and successively transmitted cases in clusters. (D) Age distribution by clinical severity. (E) Age distribution of contacts. (F) Age distributions of COVID-19 cases by type of exposure.*

### Characteristics of clusters of COVID-19

Cluster size was defined as the total number of *COVID-19 symptomatic cases* and asymptomatic individuals in a cluster. We characterized 123 clusters with clear evidence of human-to-human transmission, which includes 499 of the COVID-19 cases presented in Tab. S2. Cluster size distribution was bimodal, with most clusters were between 2 and 4 cases (94/123, corresponding to 76.4%). The largest cluster included 20 cases. The median cluster size was 3 (Tab. S2).

***Table S2.*** *Characteristics of cluster with human-to-human SARS-CoV-2 transmission identified in Hunan province, China.*

| | Number of clusters of a given size [a] (n=123) | Total number of cases (n=499) | Index case (n=142) | Case with single exposure | | | Cases with multiple exposures (n=51) | Number of infectors with a given number of transmission events [c] |
|---|---|---|---|---|---|---|---|---|
| | | | | Secondary case (n=234) | Third-generation case (n=59) | Fourth-generation case (n=13) | | |
| 0 case | - | - | - | - | - | - | - | 214 (42.9) |
| 1 case | - | - | - | - | - | - | - | 109 (21.8) |
| 2 cases | 45 (36.6) | 90 (18.0) | 45 (31.7) | 45 (19.2) | 0 (0) | 0 (0) | 0 (0) | 39 (7.8) |
| 3 cases | 26 (21.1) | 78 (15.6) | 34 (23.9) | 38 (16.2) | 0 (0) | 0 (0) | 6 (11.8) | 15 (3.0) |
| 4 cases | 23 (18.7) | 92 (18.4) | 29 (20.4) | 42 (17.9) | 8 (13.6) | 0 (0) | 13 (25.5) | 8 (1.6) |
| 5 cases | 10 (8.1) | 50 (10.0) | 11 (7.7) | 27 (11.5) | 6 (10.2) | 1 (7.7) | 5 (9.8) | 1 (0.2) |
| >5 cases | 19 (15.4) | 189 (37.9) | 23 (16.2) | 82 (35.0) | 45 (76.3) | 12 (92.3) | 27 (52.9) | 5 (1.0) |

a. Index cases were included when calculating the clusters size for each cluster.

b. Number of clustering cases at a given size of group.

c. Infectors include those with single and multiple exposures.

### Incubation period

We estimated the time from infection to symptom onset (i.e., the incubation period) based on information about the likely exposure of confirmed COVID-19 cases. Only cluster cases with confirmed human-to-human transmission and no travel history to Wuhan/Hubei were included for estimation. The rationale for this choice is that in multiple circumstances entire clusters took part in the same trip to/from Wuhan, thus preventing the unambiguous identification of the source of infection and transmission chain. Therefore, to provide more robust estimates and avoid multiplicity of biases, we have filtered those clusters. The exposure information was provided in the form of a time interval bounded by the dates of the first and last possible exposure. If the exposure start date of the case was missing or before that of the first infector, it was replaced by the exposure start date of the first infector. For the rest cases without dates of first exposure (17 individuals), they were imputed by the random numbers generated from a gamma distribution that best fitted the data of time intervals between the first and last exposure. As a sensitivity analysis, first exposure date of 7 individuals was imputed using the date when their infector came back to Hunan from Wuhan. Another sensitivity analysis was performed by excluding these 17 cases. We estimated the distribution of interval-censored exposure data by using maximum likelihood and compared three distributions (Weibull, gamma, and lognormal). The goodness of fit was assessed using Akaike information criterion (AIC). Results are presented in Tab. S3.

***Table S3.*** *Estimates of the incubation period based on the analysis of 114 clusters and 268 cases.*

| Distribution | Parameters [mean (SD)] | Mean (days) | Quantiles (0.025-0.975, days) | AIC |
|---|---|---|---|---|
| **Gamma** | shape = 2.08(0.21), rate = 0.33 (0.04) | 6.3 | 0.8 – 17.7 | 782.6 |
| **Weibull** | shape = 1.58 (0.09), scale=7.11(0.33) | 6.4 | 0.7 – 16.6 | 775.5 |
| **Lognormal** | meanlog = 1.57(0.06), sdlog = 0.82(0.04) | 6.7 | 1.0 – 23.9 | 839.7 |
| **Sensitivity analysis[a]** | | | | |
| **Gamma** | shape = 2.05(0.22), rate = 0.33 (0.04) | 6.2 | 0.7 – 17.4 | 751.3 |
| **Weibull** | shape = 1.57 (0.09), scale=6.95(0.33) | 6.3 | 0.6 – 16.3 | 744.4 |
| **lognormal** | meanlog = 1.55(0.06), sdlog = 0.83(0.04) | 6.6 | 0.9 – 23.6 | 806.7 |
| **Sensitivity analysis[b]** | | | | |
| **Gamma** | shape = 2.05 (0.22), rate = 0.33 (0.04) | 6.1 | 0.7 – 17.1 | 733.6 |
| **Weibull** | shape = 1.57 (0.09), scale=6.85(0.33) | 6.2 | 0.6 – 16.1 | 726.8 |
| **lognormal** | meanlog = 1.53(0.06), sdlog = 0.83(0.05) | 6.4 | 0.9 – 23.3 | 788.4 |

a. Sensitivity analysis performed based on 258 cases including 7 individuals for which the first exposure date was imputed using the date when their infector came back to Hunan from Wuhan.
b. Sensitivity analysis performed based on 251 cases (i.e., excluding 17 individuals without first exposure date).

### Serial interval

We analyzed clusters of COVID-19 cases with known epidemiological links and no travel history to Wuhan/Hubei to estimate the interval between onset of symptoms in primary (index) cases and the onset of symptoms in secondary cases generated by these primary cases (i.e., the serial intervals). For cases with several possible infectors, a time interval bounded by the symptom-onset dates of the first and last possible infectors was provided as the symptom onset interval of primary cases. Using dates of symptom onsets for consecutive generations of cases within clusters, we fitted a gamma distribution with a shift parameter allowing negative serial intervals of interval-censored data by maximum likelihood to estimate the distribution of serial interval. The epidemic was further divided into two time periods (January 5 to January 23, and January 24 to April 2) by using the date of symptom onset relative to the date of level 1 emergency response activation in Hunan province (January 24). The overall and phased-in estimation of serial intervals are presented in main text and Tab. S4. Only the transmission pairs with a unanimously identified infector were used in this analysis.

***Table S4.*** *Estimates of serial interval based on the analysis of 245 transmission pairs.*

| **Period*** | **Sample size** | **Parameters [mean (SD)]** | **Mean (days)** | **IQR (0.025-0.975, days)** |
|---|---|---|---|---|
| Overall | 245 | shape = 9.68(0.86), rate = 0.48(0.04), shift=14.5 | 5.5 | -5.0 – 19.9 |
| January 5 - January 23 | 111 | shape = 19.07(2.54), rate = 0.88(0.12), shift=14.5 | 7.0 | -1.5 - 17.7 |
| January 24 - April 2 | 110 | shape = 6.59(0.87), rate = 0.35(0.05), shift=14.5 | 4.1 | -7.3 – 20.9 |

*Period was defined using the date of symptom onset of the infector in each transmission pair.

### Infectiousness profile over time

Following the approach similar to He, et al [2], and accounting for the correction proposed by

Ashcroft, et al [3], the infectiousness profile (i.e., transmission probability from primary cases to a secondary case) was inferred using the serial intervals from confirmed transmission pairs combined with the incubation period distribution fitted in our analysis. Assuming that the infectiousness profile $\beta_c(t_I - t_{S1})$ follows a gamma distribution with a time shift c to allow for start of infectiousness ($t_I$) c days prior to the date of symptom onset ($t_{S1}$). The serial intervals distribution $f(t_{S2} - t_{S1})$ would be the convolution between the infectiousness profile and incubation period distribution $g(t_{S2} - t_I)$, where $t_{S2}$ is the date when secondary case shows symptoms. The parameter vector θ, which includes shape and scale of the gamma distribution and the time shift c, were estimated using maximum likelihood based on the convolution of serial interval and incubation period. Allowing for the start of infectiousness to be around symptom onset and taking into account the window of symptom onset ($t_{S1l}, t_{S1u}$), the likelihood function was given by

$$L(t_{S1u}, t_{S1l,} t_{S2} | \theta) = \int_{t_{S1l,}}^{t_{S1u}} \int_{-\infty}^{t_{S2}} \beta_c(t_I - t_{S1}) g(t_{S2} - t_I) d_{t_I} d_{t_{S1}}$$

The results of the estimation are presented in the main text.

### Generation time

Generation time - that is the time interval between infection of the primary case ($t_{I1}$) and infection of the secondary cases ($t_{I2}$) generated by such primary case - was inferred using the data of incubation period combined with infectiousness profile estimated in our analysis. We considered that infected cases would show symptoms at certain time ($t_S$) before or after onset of infectiousness. Assuming that the distribution of generation time follows a gamma distribution $\varphi(t_{I2} - t_{I1})$, the observed distribution of incubation period $g(t_S - t_{I1})$ can be inferred as the convolution between the infectiousness profile $\beta_c(t_{I2} - t_S)$ and the generation time distribution. We constructed a likelihood function based on the convolution, which was fitted to the observed incubation period, with $t_{I1}$ provided in the form of a time interval bounded by the dates of the first and last possible exposure ($t_{E1}$, $t_{E2}$), given by

$$L(t_{E1}, t_{E2}, t_S|\alpha, \beta) = \int_{t_{E1}}^{t_{E2}} \int_{t_{I1}}^{+\infty} \varphi(t_{I2} - t_{I1})\beta_c(t_{I2} - t_S) d_{t_S} d_{t_{I1}}$$

Shape parameter ($\alpha$) and rate parameter ($\beta$) of the gamma distribution of generation time were estimated using maximum likelihood method. The generation time was estimated to be 5.7 days (median: 5.5 days, interquartile range: 4.5, 6.7 days) based on a gamma distribution (shape=10.56, rate=1.85).

#### Other key time-to-event intervals

Other key time-to-event distributions were estimated by using maximum likelihood. In particular, we estimated: i) the time from symptom onset to the date of collection of the first sample for PCR testing and ii) the time from symptom onset to laboratory confirmation. Three distributions (Weibull, gamma, and lognormal) with shift parameters allowing negative intervals were fitted and compared. The goodness of fit was assessed using AIC.

As described above, the infectiousness profile peaked before the day of symptom onset. This may be driven by the control measures like isolation of infectors. We estimated the distribution of interval from symptom onset to the sampling date of first PCR and to laboratory confirmation to evaluate the timing of identification, isolation, and diagnosis of infectious individuals. Results are presented in the main text and Tab. S5 (where only the best fitting distribution is shown).

***Table S5.*** *Estimates of other key time-to-event intervals*

| Best fitted distribution | Sample size | Parameters [mean (SD)] | Mean (days) | quantiles (0.025-0.975, days) |
|---|---|---|---|---|
| **Time from symptom onset to date of collection of the first sample for PCR testing** | | | | |
| Gamma | 531 | shape = 12.05(0.73), rate = 0.76(0.05), shift=11.1 | 4.7 | -3.0-14.8 |
| **Time from symptom onset to laboratory confirmation** | | | | |
| **Gamma** | 952 | shape = 9.45(0.43), rate = 0.70(0.03), shift=7 | 6.4 | -7.2 – 16.3 |

### Clusters shown evidence for asymptomatic transmission

From the analysis of contact tracing records, we identified 8 clusters with evidence of asymptomatic transmission as shown in Fig. S5.

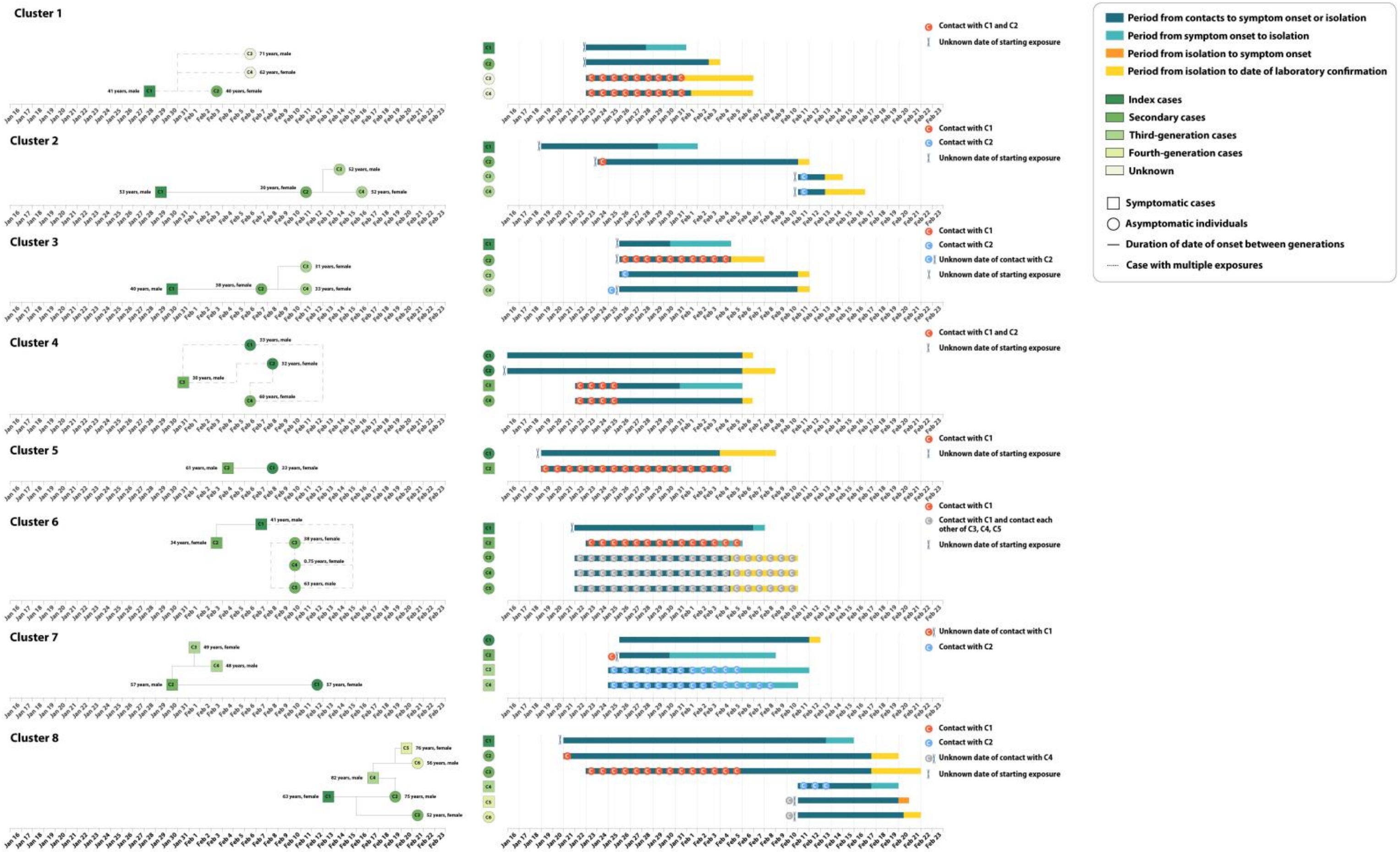

***Figure. S5.*** *Transmission chain in all the clusters showing evidence of asymptomatic SARS-CoV-2 transmission. Square symbols indicate symptomatic cases and circular symbols indicate asymptomatic subjects. Age, sex and generation in a cluster are shown for each SARS-CoV-2 infected individual (left panels), with information on date of diagnosis to the first RT-PCR positive for asymptomatic subjects. Timeline of events (right panels).*

### Total and mean number of infections by age of infector and of infectee

From 254 certain transmission pairs, we estimate the total (Fig. S6A) and mean (Fig. S6B) number of infections by age. These matrices are descriptive and do not account for confounding factors other than age. Therefore, they cannot be used to estimate susceptibility and infectivity by age group. For example, the lower mean number of infections generated by children (0-14 years old) with respect to adults is the joint effect of several factors. According to our regression analysis, one of these factors is the generation of infection. Infected individuals in generation one have much higher odds of transmitting the infection, probably due to the case isolation and quarantine of close contacts that increase with the generation. Coupled with the low proportion of children in the first generation as compared to adults (we remind that the schools were closed during the entire study period and close community management policies were in place), this may have contributed to lower number of infections generated by children. The summary tables by age and generation are reported in Tab. S6 and Tab. S7.

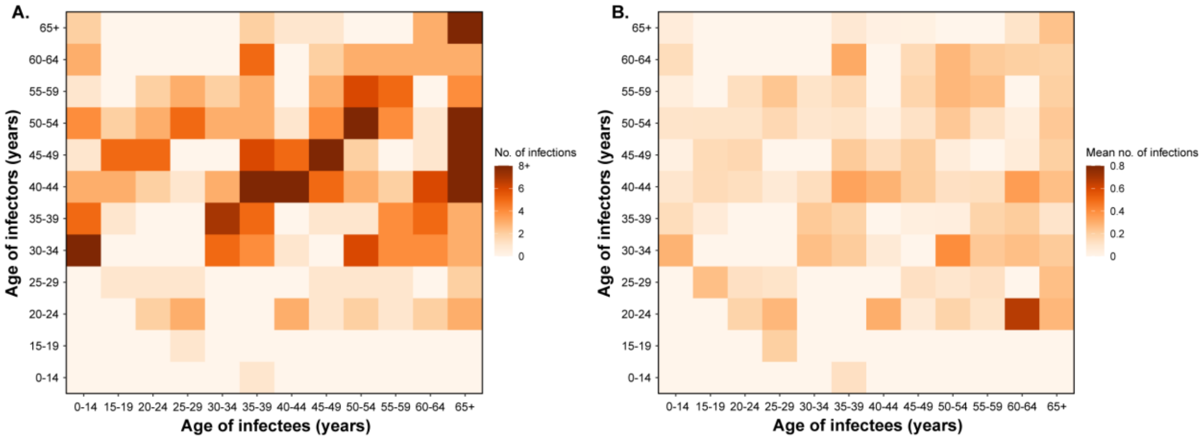

***Figure S6.*** *Number of infections by age of infector and of infectee. Each cell in the matrices refers to the total number of infections (A) and the mean number of infections (B) caused by an infector of a given age.*

***Table S6.*** *Summary of contact tracing data by age of infectors and generation of transmission.*

| | No. of infectors | Number of infected contacts (independent of contact's age) | Number of contacts (independent of contact's age) | % of infected among contacts (N of infected contacts/ N of contacts) | Average number of infector's contacts |
|---|---|---|---|---|---|
| Age of infectors | | | | | |
| 0-14 y | 25 | 2 | 193 | 1.04 | 7.72 |
| G1 | 1 | 0 | 1 | 0 | - |
| G2 | 7 | 1 | 59 | 1.69 | - |
| G3-4 | 6 | 0 | 52 | 0 | - |
| Multiple exposure | 3 | 1 | 25 | 4.00 | - |
| Unknown | 8 | 0 | 56 | 0 | - |
| 15-64 y | 355 | 188 | 6,833 | 3.00 | 19.25 |
| G1 | 59 | 84 | 1,951 | 4.00 | - |
| G2 | 149 | 70 | 2,324 | 3.00 | - |
| G3-4 | 50 | 17 | 839 | 2.03 | - |
| Multiple exposure | 30 | 8 | 551 | 1.45 | - |
| Unknown | 67 | 9 | 1,168 | 1.00 | - |
| 65+ y | 81 | 19 | 1,133 | 2.00 | 13.99 |
| G1 | 6 | 7 | 169 | 4.14 | - |

| | | | | | |
|---|---|---|---|---|---|
| G2 | 46 | 10 | 604 | 1.66 | - |
| G3-4 | 7 | 2 | 74 | 2.70 | - |
| Multiple exposure | 4 | 0 | 22 | 0 | - |
| Unknown | 18 | 0 | 264 | 0 | - |

***Table S7.*** *Summary of contact tracing data by age of contacts and generation of transmission.*

| | Number of contacts | Number of infected contacts | Total number of infectors (independent of infector's age) | % of infected among contacts (N of infected contacts/ N of contacts) | Average number of infector's contacts |
|---|---|---|---|---|---|
| Age of contacts | | | | | |
| 0-14 y | 936 | 22 | 309 | 2.35 | 3.03 |
| G1 | 174 | 7 | 50 | 4.02 | - |
| G2 | 349 | 8 | 131 | 2.29 | - |
| G3-4 | 131 | 5 | 46 | 3.82 | - |
| Multiple exposure | 82 | 1 | 26 | 1.22 | - |
| Unknown | 200 | 1 | 56 | 0.50 | - |
| 15-64 y | 6,411 | 154 | 446 | 2.00 | 14.37 |
| G1 | 1,723 | 65 | 66 | 4.00 | - |
| G2 | 2,363 | 60 | 198 | 3.00 | - |
| G3-4 | 718 | 13 | 61 | 1.81 | - |
| Multiple exposure | 466 | 8 | 33 | 1.72 | - |
| Unknown | 1,141 | 8 | 88 | 1.00 | - |
| 65+ y | 812 | 33 | 279 | 4.06 | 2.91 |
| G1 | 224 | 19 | 50 | 8.48 | - |
| G2 | 275 | 13 | 114 | 4.73 | - |
| G3-4 | 116 | 1 | 41 | 0.86 | - |
| Multiple exposure | 50 | 0 | 19 | 0 | - |
| Unknown | 147 | 0 | 55 | 0 | - |

### Descriptive univariate analysis

To describe the correlations between the single factors and the probability of successful/unsuccessful transmission, we performed a univariate generalized linear model analysis. The results are presented in Tab. S8. It is important to note that this analysis does not account for the confounding effect of multiple factors and thus does not provide reliable estimates for the inference of the effect of the covariates and their statistical significance. For this reason, in the following section, we performed a multivariate analysis.

***Table S8.*** *Univariate analysis of factors possibly connected with SARS-CoV-2 transmission patters.*

| Characteristics | No. of contact | Univariate analysis | |
|---|---|---|---|
| | | OR (95%CI) | P-value |
| Age of infectors | | | |
| 0-14 y | 193 | 0.33 (0.04, 2.83) | 0.313 |
| 15-64 y | 6,833 | Reference | - |
| 65+ y | 1,133 | 0.59 (0.21, 1.65) | 0.316 |
| Log-transformed age | 8,159 | 1.75 (0.87, 3.54) | 0.120 |
| Age of contacts | | | |
| 0-14 y | 936 | 0.79 (0.47, 1.32) | 0.371 |
| 15-64 y | 6411 | Reference | - |
| 65+ y | 812 | 1.82 (1.15, 2.86) | 0.010 |
| Log-transformed age | 8,159 | 1.14 (0.93, 1.40) | 0.196 |
| Type of contact | | | |
| Household contacts | 1,021 | Reference | - |
| Relative contacts | 3,084 | 0.12 (0.08, 0.19) | <0.001 |
| Social contacts | 2,227 | 0.06 (0.04, 0.12) | <0.001 |
| Other contacts | 1,827 | 0.08 (0.04, 0.14) | <0.001 |
| Generation of SARS-CoV-2 transmission | | | |
| G1 | 2,121 | Reference | - |
| G2 | 2,987 | 0.15 (0.06, 0.39) | <0.001 |
| G3-4 | 965 | 0.13 (0.03, 0.49) | 0.003 |
| Multiple exposure[a] | 598 | 0.13 (0.03, 0.64) | 0.012 |
| Unknown | 1,488 | 0.03 (0.01, 0.12) | <0.001 |
| Levels of exposure to an infector | | | |
| Total number of contacts | 8,159 | 0.99 (0.97, 1.00) | 0.060 |
| Gender of infectors | | | |
| Female | 4,067 | Reference | - |
| Male | 4,092 | 1.92 (0.93, 3.94) | 0.077 |
| Gender of contacts | | | |
| Female | 4,017 | Reference | - |
| Male | 4,142 | 0.93 (0.68, 1.26) | 0.628 |
| Clinical severity of infectors | | | |
| Asymptomatic subjects | 898 | 0.53 (0.18, 1.56) | 0.247 |
| Symptomatic subjects | 7,261 | Reference | - |

[a] Contacts who were exposed to multiple cases of different generations of SARS-CoV-2 transmission.

### Quantifying the impact of potential drivers on the susceptibility and infectivity of

#### SARS-CoV-2

We analyzed the odds ratio of SARS-CoV-2 transmission given the characteristics of the infectors and their contacts. To consider the clustering effect of an infector and a cluster, mixed effect logit models (i.e., generalized linear mixed-effect model, GLMM, for binary data with the logit link) were used to explore potential drivers of the susceptibility and infectivity of SARS-CoV-2 virus. The specifications of the GLMM models are defined as follows:

$$g(u_i) = \alpha + \beta_1 Age\_infector_i + \beta_2 Age\_contact_i + \beta_3 Contact\_type_i + \beta_4 Generation\_infector_i + \beta_5 Exposure\_level_i + \beta_6 Gender\_infector_i + \beta_7 Gender\_contact_i + \beta_8 Case\_type_i + \beta_9 Observation\_period_i + u_0 + u_1$$

Where:

- g is a logit link function;
- $\alpha$ is the intersect
- $Age\_infector_i$ is the fixed effects of the age group of the infector in the successful (1) or unsuccessful (0) transmission event $i$;
- $Age\_contact_i$ is the age group of the contact (potential infectee) in the successful/unsuccessful transmission event $i$;
- $Contact\_type_i$ is the type of contact occurred in the successful/unsuccessful transmission event $i$;
- $Generation\_infector_i$ is the generation of the successful/unsuccessful transmission event $i$;
- $Exposure\_level_i$ is the number of close contacts of the infector involved in the successful/unsuccessful transmission event $i$;

- $Gender\_infector_i$ is the gender of the infector in the successful/unsuccessful transmission event $i$;
- $Gender\_contact_i$ is the gender of contact in the successful/unsuccessful transmission event $i$;
- $Case\_type_i$ discriminates whether the infector involved in the successful/unsuccessful transmission event $i$ is symptomatic or asymptomatic;
- $Observation\_period_i$ indicates the observation period for an infector/contact involved in the successful/unsuccessful transmission event $i$;
- $u_0$ and $u_1$ are random effects attributed to an infector and a cluster, respectively. $u_i = E[Y|(u_0, u_1)]$ is the mean of the response variable $Y_i$ of a given value of the random effects.

The results of the multivariate analysis based on GLMM are presented in Table S9. The results for fixed effects, including 3 age groups for infector's and infectee's age, are presented in the Table S10 and Figure S8. To evaluate the disaggregated effects of age, we also used transformed (log) continuous age variables (i.e., age of infectors and contacts) (Tab. S11). The goodness-of-fit evaluation was based on the estimates provided in the Table S12. Model diagnostic measures and residuals plots (Fig. S7) were evaluated by DHARMa residual diagnostics for hierarchical models [4].

To further explore how the probability of SARS-COV-2 infections changes with a change in each covariate, the average marginal effects of age of infector and contacts, type of contact between infector and contact were estimated across all contacts, holding the effect of other covariate constant (Fig. S9).

In addition, to explore possible non-linearity in the connection of age and of the number of contacts with SARS-CoV-2 transmission, we used generalized additive mixed models (GAMM). We used the same specifications as in the GLMM models presented in the main text. The summary of the results of the GAMM models is shown in Fig. S10 and in Tab. S13. The obtained results suggest that the risk of SARS-CoV-2 transmission monotonically increase with the age of contacts and with the number of infector's contacts. This is consistent with the patterns that have been shown in GLMM models.

To explore the possible effect of timing on the results of the regression analysis we have also introduced an additional variable identifying the three phases of the epidemic: 1. Before the level 1 emergency response was activated in Hunan province (Jan 24); 2. After the level 1 emergency response activation, but before the growth of cases was reversed (Fig. S2); 3. After the outbreak growth was reversed.

***Table S9.*** *Random effect of infectors and clusters in generalized linear mixed model with categorized age of infectors and contacts.*

| | Infector | | Cluster | |
|---|---|---|---|---|
| | Variance | Standard deviation | Variance | Standard deviation |
| Step 1-1 | 6.14 | 2.48 | 0.27 | 0.52 |
| Step 1-2 | 3.88 | 1.97 | 0.75 | 0.87 |
| Step 1-3 | 3.06 | 1.75 | 0.99 | 1.00 |
| Step 1-4 | 2.98 | 1.73 | 0.96 | 0.98 |
| Step 1-5 | 2.98 | 1.73 | 0.96 | 0.98 |
| Step 1-6[a] | 2.88 | 1.70 | 1.04 | 1.02 |
| Step 1-7 | 2.78 | 1.67 | 0.95 | 0.98 |
| Step 2-1 | 5.98 | 2.45 | 0.34 | 0.58 |
| Step 2-2 | 3.73 | 1.93 | 0.84 | 0.92 |

| | | | | |
|---|---|---|---|---|
| Step 2-3 | 3.01 | 1.74 | 1.06 | 1.03 |
| Step 2-4 | 2.94 | 1.71 | 1.01 | 1.00 |
| Step 2-5 | 2.94 | 1.71 | 1.01 | 1.00 |
| Step 2-6[b] | 2.85 | 1.69 | 1.08 | 1.04 |
| Step 2-7 | 2.78 | 1.67 | 0.97 | 0.98 |

[a] model 1

[b] model 2

***Table S10.*** *Stepwise regression analysis of factors associated with the probability of acquiring SARS-CoV-2 infections in generalized linear mixed models with categorized age of infectors and contacts.*

| Characteristics | No. of contact | Step 1-1 | | Step 1-2 | | Step 1-3 | | Step 1-4 | | Step 1-5 | | Step 1-6 [a] | | Step 1-7 | |
|---|---|---|---|---|---|---|---|---|---|---|---|---|---|---|---|
| | | OR (95%CI) | P-value | OR (95%CI) | P-value | OR (95%CI) | P-value | OR (95%CI) | P-value | OR (95%CI) | P-value | OR (95%CI) | P-value | OR (95%CI) | P-value |
| Intercept | - | 0.03 (0.01, 0.06) | <0.001 | 0.22 (0.11, 0.44) | <0.001 | 0.37 (0.17, 0.83) | 0.015 | 0.27 (0.11, 0.65) | 0.003 | 0.27 (0.11, 0.65) | 0.004 | 0.28 (0.12, 0.68) | 0.005 | 0.23 (0.09, 0.57) | 0.001 |
| Age of infectors | | | | | | | | | | | | | | | |
| 0-14 y | 193 | 0.17 (0.02, 1.35) | 0.093 | 0.28 (0.04, 2.18) | 0.226 | 0.24 (0.03, 1.72) | 0.155 | 0.25 (0.04, 1.75) | 0.161 | 0.25 (0.04, 1.75) | 0.162 | 0.28 (0.04, 2.04) | 0.210 | 0.27 (0.04, 1.89) | 0.185 |
| 15-64 y | 6,833 | Reference | - | Reference | - | Reference | - | Reference | - | Reference | - | Reference | - | Reference | - |
| 65+ y | 1,133 | 0.46 (0.17, 1.25) | 0.128 | 0.65 (0.25, 1.71) | 0.382 | 0.63 (0.25, 1.58) | 0.326 | 0.64 (0.26, 1.59) | 0.332 | 0.64 (0.26, 1.59) | 0.332 | 0.62 (0.25, 1.55) | 0.306 | 0.61 (0.25, 1.50) | 0.280 |
| Age of contacts | | | | | | | | | | | | | | | |
| 0-14 y | 936 | 0.55 (0.32, 0.93) | 0.026 | 0.58 (0.34, 0.98) | 0.043 | 0.58 (0.34, 0.98) | 0.041 | 0.58 (0.34, 0.98) | 0.042 | 0.58 (0.34, 0.98) | 0.042 | 0.58 (0.34, 0.98) | 0.041 | 0.58 (0.34, 0.98) | 0.042 |
| 15-64 y | 6,411 | Reference | - | Reference | - | Reference | - | Reference | - | Reference | - | Reference | - | Reference | - |
| 65+ y | 812 | 1.66 (1.03, 2.67) | 0.038 | 1.66 (1.03, 2.68) | 0.036 | 1.64 (1.02, 2.62) | 0.041 | 1.65 (1.03, 2.65) | 0.038 | 1.65 (1.03, 2.65) | 0.038 | 1.65 (1.03, 2.64) | 0.038 | 1.65 (1.03, 2.65) | 0.037 |
| Type of contact | | | | | | | | | | | | | | | |
| Household contacts | 1,021 | Reference | - | Reference | - | Reference | - | Reference | - | Reference | - | Reference | - | Reference | - |
| Relative contacts | 3,084 | 0.11 (0.07, 0.17) | <0.001 | 0.10 (0.07, 0.16) | <0.001 | 0.11 (0.07, 0.17) | <0.001 | 0.11 (0.07, 0.17) | <0.001 | 0.11 (0.07, 0.17) | <0.001 | 0.11 (0.07, 0.17) | <0.001 | 0.11 (0.07, 0.18) | <0.001 |
| Social contacts | 2,227 | 0.06 (0.03, 0.11) | <0.001 | 0.05 (0.03, 0.10 | <0.001 | 0.06 (0.03, 0.11) | <0.001 | 0.06 (0.03, 0.11) | <0.001 | 0.06 (0.03, 0.11) | <0.001 | 0.06 (0.03, 0.11) | <0.001 | 0.06 (0.03, 0.11) | <0.001 |
| Other contacts | 1,827 | 0.07 (0.04, 0.13) | <0.001 | 0.07 (0.04, 0.12) | <0.001 | 0.07 (0.04, 0.13) | <0.001 | 0.07 (0.04, 0.13) | <0.001 | 0.07 (0.04, 0.13) | <0.001 | 0.07 (0.04, 0.13) | <0.001 | 0.07 (0.04, 0.13) | <0.001 |
| Generation of SARS-CoV-2 transmission | | | | | | | | | | | | | | | |
| G1 | 2,121 | - | - | Reference | - | Reference | - | Reference | - | Reference | - | Reference | - | Reference | - |
| G2 | 2,987 | - | - | 0.15 (0.06, 0.36) | <0.001 | 0.13 (0.05, 0.29) | <0.001 | 0.13 (0.06, 0.31) | <0.001 | 0.13 (0.06, 0.31) | <0.001 | 0.14 (0.06, 0.32) | <0.001 | 0.14 (0.06, 0.32) | <0.001 |
| G3-4 | 965 | - | - | 0.07 (0.02, 0.27) | <0.001 | 0.05 (0.02, 0.19) | <0.001 | 0.05 (0.02, 0.19) | <0.001 | 0.05 (0.02, 0.19) | <0.001 | 0.05 (0.02, 0.19) | <0.001 | 0.06 (0.02, 0.21) | <0.001 |
| Multiple exposure[b] | 598 | - | - | 0.11 (0.03, 0.47) | 0.003 | 0.09 (0.02, 0.35) | <0.001 | 0.10 (0.03, 0.41) | 0.001 | 0.10 (0.03, 0.41) | 0.001 | 0.11 (0.03, 0.43) | 0.002 | 0.11 (0.03, 0.43) | 0.001 |
| Unknown | 1,488 | - | - | 0.03 (0.01, 0.11) | <0.001 | 0.03 (0.01, 0.10) | <0.001 | 0.03 (0.01, 0.1) | <0.001 | 0.03 (0.01, 0.10) | <0.001 | 0.03 (0.01, 0.11) | <0.001 | 0.03 (0.01, 0.10) | <0.001 |
| Levels of exposure to an infector | | | | | | | | | | | | | | | |
| Total number of contacts | 8,159 | - | - | - | - | 0.99 (0.97, 1.00) | 0.028 | 0.99 (0.97, 1.00) | 0.028 | 0.99 (0.97, 1.00) | 0.028 | 0.99 (0.97, 1.00) | 0.022 | 0.99 (0.97, 1.00) | 0.043 |
| Gender of infectors | | | | | | | | | | | | | | | |
| Female | 4,067 | - | - | - | - | - | - | Reference | - | Reference | - | Reference | - | Reference | - |
| Male | 4,092 | - | - | - | - | - | - | 1.76 (0.97, 3.21) | 0.064 | 1.76 (0.97, 3.21) | 0.063 | 1.75 (0.96, 3.17) | 0.067 | 1.66 (0.92, 3.00) | 0.096 |

| | | | | | | | | | | | | | | | |
|---|---|---|---|---|---|---|---|---|---|---|---|---|---|---|---|
| Gender of contacts | | | | | | | | - | - | | | | | | |
| Female | 4,017 | - | - | - | - | - | - | - | - | Reference | - | Reference | - | Reference | - |
| Male | 4,142 | - | - | - | - | - | - | - | - | 1.02 (0.74, 1.40) | 0.907 | 1.02 (0.74, 1.41) | 0.899 | 1.02 (0.74, 1.40) | 0.902 |
| Clinical severity of infectors | | | | | | | | | | | | | | | |
| Asymptomatic subjects | 898 | - | - | - | - | - | - | - | - | - | - | 0.68 (0.25, 1.88) | 0.460 | 0.58 (0.21, 1.61) | 0.293 |
| Symptomatic subjects | 7,261 | - | - | - | - | - | - | - | - | - | - | Reference | - | Reference | - |
| Period of observation[c] | | | | | | | | | | | | | | | |
| Before January 23, 2020 | 198 | - | - | - | - | - | - | - | - | - | - | - | - | 2.98 (0.46, 19.31) | 0.253 |
| January 24, 2020- February 4, 2020 | 5,272 | - | - | - | - | - | - | - | - | - | - | - | - | Reference | - |
| After February 4, 2020 | 2,689 | - | - | - | - | - | - | - | - | - | - | - | - | 1.72 (0.84, 3.51) | 0.137 |

[a] model 1

[b] Contacts who were exposed to multiple cases of different generations of SARS-CoV-2 transmission.

[c] Referring to three phases of epidemic control and major changes in COVID-19 case definition, period of observation was defined using quarantine and isolation date, as well as date of diagnosis.

***Table S11.*** *Stepwise regression analysis of factors associated with the probability of acquiring SARS-CoV-2 infections in generalized linear mixed models with log-transformed age of infectors and contacts.*

| Characteristics | No. of contact | Step 2-1 | | Step 2-2 | | Step 2-3 | | Step 2-4 | | Step 2-5 | | Step 2-6[a] | | Step 2-7 | |
|---|---|---|---|---|---|---|---|---|---|---|---|---|---|---|---|
| | | OR (95%CI) | P-value | OR (95%CI) | P-value | OR (95%CI) | P-value | OR (95%CI) | P-value | OR (95%CI) | P-value | OR (95%CI) | P-value | OR (95%CI) | P-value |
| Intercept | - | 0 (0, 0.01) | <0.001 | 0.01 (0, 0.19) | 0.001 | 0.02 (0, 0.29) | 0.003 | 0.02 (0, 0.21) | 0.002 | 0.02 (0, 0.21) | 0.002 | 0.02 (0, 0.26) | 0.003 | 0.02 (0, 0.21) | 0.002 |
| Age of infectors | | | | | | | | | | | | | | | |
| Log-transformed age | 8,159 | 1.92 (0.99, 3.73) | 0.054 | 1.62 (0.87, 3.01) | 0.128 | 1.61 (0.90, 2.90) | 0.108 | 1.62 (0.91, 2.89) | 0.103 | 1.62 (0.91, 2.90) | 0.104 | 1.57 (0.87, 2.81) | 0.134 | 1.56 (0.88, 2.77) | 0.130 |
| Age of contacts | | | | | | | | | | | | | | | |
| Log-transformed age | 8,159 | 1.28 (1.04, 1.58) | 0.019 | 1.26 (1.03, 1.56) | 0.028 | 1.26 (1.02, 1.55) | 0.029 | 1.26 (1.02, 1.55) | 0.028 | 1.26 (1.02, 1.55) | 0.028 | 1.26 (1.02, 1.55) | 0.028 | 1.26 (1.02, 1.55) | 0.028 |
| Type of contact | | | | | | | | | | | | | | | |
| Household contacts | 1,021 | Reference | - | Reference | - | Reference | - | Reference | - | Reference | - | Reference | - | Reference | - |

| | | | | | | | | | | | | | | | |
|---|---|---|---|---|---|---|---|---|---|---|---|---|---|---|---|
| Relative contacts | 3,084 | 0.11 (0.07, 0.18) | <0.001 | 0.11 (0.07, 0.17) | <0.001 | 0.11 (0.07, 0.18) | <0.001 | 0.11 (0.07, 0.18) | <0.001 | 0.11 (0.07, 0.18) | <0.001 | 0.11 (0.07, 0.18) | <0.001 | 0.12 (0.07, 0.18) | <0.001 |
| Social contacts | 2,227 | 0.06 (0.03, 0.11) | <0.001 | 0.05 (0.03, 0.09) | <0.001 | 0.06 (0.03, 0.11) | <0.001 | 0.06 (0.03, 0.11) | <0.001 | 0.06 (0.03, 0.11) | <0.001 | 0.06 (0.03, 0.11) | <0.001 | 0.06 (0.03, 0.10) | <0.001 |
| Other contacts | 1,827 | 0.07 (0.04, 0.13) | <0.001 | 0.07 (0.04, 0.12) | <0.001 | 0.07 (0.04, 0.13) | <0.001 | 0.07 (0.04, 0.13) | <0.001 | 0.07 (0.04, 0.13) | <0.001 | 0.07 (0.04, 0.13) | <0.001 | 0.07 (0.04, 0.13) | <0.001 |
| Generation of SARS-CoV-2 transmission | | | | | | | | | | | | | | | |
| G1 | 2,121 | - | - | Reference | - | Reference | - | Reference | - | Reference | - | Reference | - | Reference | - |
| G2 | 2,987 | - | - | 0.14 (0.06, 0.33) | <0.001 | 0.12 (0.05, 0.27) | <0.001 | 0.12 (0.05, 0.28) | <0.001 | 0.12 (0.05, 0.28) | <0.001 | 0.13 (0.05, 0.29) | <0.001 | 0.13 (0.06, 0.30) | <0.001 |
| G3-4 | 965 | - | - | 0.07 (0.02, 0.25) | <0.001 | 0.05 (0.01, 0.18) | <0.001 | 0.05 (0.02, 0.18) | <0.001 | 0.05 (0.02, 0.18) | <0.001 | 0.05 (0.02, 0.18) | <0.001 | 0.06 (0.02, 0.20) | <0.001 |
| Multiple exposure[b] | 598 | - | - | 0.11 (0.03, 0.44) | 0.002 | 0.08 (0.02, 0.34) | <0.001 | 0.10 (0.03, 0.40) | 0.001 | 0.10 (0.03, 0.40) | 0.001 | 0.11 (0.03, 0.42) | 0.001 | 0.11 (0.03, 0.42) | 0.001 |
| Unknown | 1,488 | - | - | 0.03 (0.01, 0.11) | <0.001 | 0.03 (0.01, 0.10) | <0.001 | 0.03 (0.01, 0.10) | <0.001 | 0.03 (0.01, 0.10) | <0.001 | 0.03 (0.01, 0.11) | <0.001 | 0.03 (0.01, 0.10) | <0.001 |
| Levels of exposure to an infector | | | | | | | | | | | | | | | |
| Total number of contacts | 8,159 | - | - | - | - | 0.99 (0.97, 1.00) | 0.037 | 0.99 (0.97, 1.00) | 0.036 | 0.99 (0.97, 1.00) | 0.036 | 0.99 (0.97, 1.00) | 0.030 | 0.99 (0.98, 1.00) | 0.057 |
| Gender of infectors | | | | | | | | | | | | | | | |
| Female | 4,067 | - | - | - | - | - | - | Reference | - | Reference | - | Reference | - | Reference | - |
| Male | 4,092 | - | - | - | - | - | - | 1.77 (0.97, 3.21) | 0.061 | 1.77 (0.97, 3.21) | 0.061 | 1.75 (0.97, 3.18) | 0.064 | 1.67 (0.92, 3.01) | 0.091 |
| Gender of contacts | | | | | | | | - | - | | | | | | |
| Female | 4,017 | - | - | - | - | - | - | - | - | Reference | - | Reference | - | Reference | - |
| Male | 4,142 | - | - | - | - | - | - | - | - | 1.01 (0.73, 1.38) | 0.973 | 1.01 (0.73, 1.39) | 0.964 | 1.01 (0.73, 1.39) | 0.968 |
| Clinical severity of infectors | | | | | | | | | | | | | | | |
| Asymptomatic subjects | 898 | - | - | - | - | - | - | - | - | - | - | 0.72 (0.26, 1.98) | 0.523 | 0.62 (0.22, 1.73) | 0.360 |
| Symptomatic subjects | 7,261 | - | - | - | - | - | - | - | - | - | - | Reference | - | Reference | - |
| Period of observation[c] | | | | | | | | | | | | | | | |
| Before January 23, 2020 | 198 | - | - | - | - | - | - | - | - | - | - | - | - | 3.15 (0.48, 20.54) | 0.230 |
| January 24, 2020- February 4, 2020 | 5,272 | - | - | - | - | - | - | - | - | - | - | - | - | Reference | - |
| After February 4, 2020 | 2,689 | - | - | - | - | - | - | - | - | - | - | - | - | 1.63 (0.80, 3.32) | 0.176 |

[a] model 2

[b] Contacts who were exposed to multiple cases of different generations of SARS-CoV-2 transmission.

[c] Referring to three phases of epidemic control and major changes in COVID-19 case definition, period of observation was defined using quarantine and isolation date, as well as date of diagnosis.

***Table S12.*** *Assessing the fit of generalized linear mixed model with categorized age of infectors and contacts.*

| Model | AIC (ΔAIC) | BIC | loglikelihood | deviance | $\chi^2$ | P-value[a] |
|---|---|---|---|---|---|---|
| Step 1-1 | 1577.5 | 1647.6 | -778.8 | 1557.5 | - | - |
| Step 1-2 | 1541.9 (35.6) | 1640.0 | -756.9 | 1513.9 | 43.612 | <0.001 |
| Step 1-3 | 1539.5 (2.4) | 1644.6 | -754.7 | 1509.5 | 4.433 | 0.035 |
| Step 1-4 | 1538.0 (1.5) | 1650.1 | -753.0 | 1506.0 | 3.456 | 0.063 |
| Step 1-5[b] | 1540.0 (-2.0) | 1659.1 | -753.0 | 1506.0 | 0.014 | 0.907 |
| Step 1-6 | 1541.4 (-1.4) | 1667.6 | -752.7 | 1505.4 | 0.553 | 0.457 |
| Step 1-7 | 1542.2 (-0.8) | 1682.4 | -751.1 | 1502.2 | 3.187 | 0.203 |
| Step 2-1 | 1579.1 | 1635.2 | -781.6 | 1563.1 | - | - |
| Step 2-2 | 1541.9 (37.2) | 1626.0 | -759.0 | 1517.9 | 45.188 | <0.001 |
| Step 2-3 | 1539.9 (2.0) | 1631.0 | -757.0 | 1513.9 | 4.024 | 0.045 |
| Step 2-4 | 1538.4 (1.5) | 1636.5 | -755.2 | 1510.4 | 3.529 | 0.060 |
| Step 2-5[c] | 1540.4 (-2.0) | 1645.5 | -755.2 | 1510.4 | 0.001 | 0.974 |
| Step 2-6 | 1542.0 (-1.6) | 1654.1 | -755.0 | 1510.0 | 0.410 | 0.522 |
| Step 2-7 | 1543.0 (-1.0) | 1669.2 | -753.5 | 1507.0 | 2.932 | 0.231 |

[a]ANOVA analyses were performed.

[b] model 1

[c] model 2

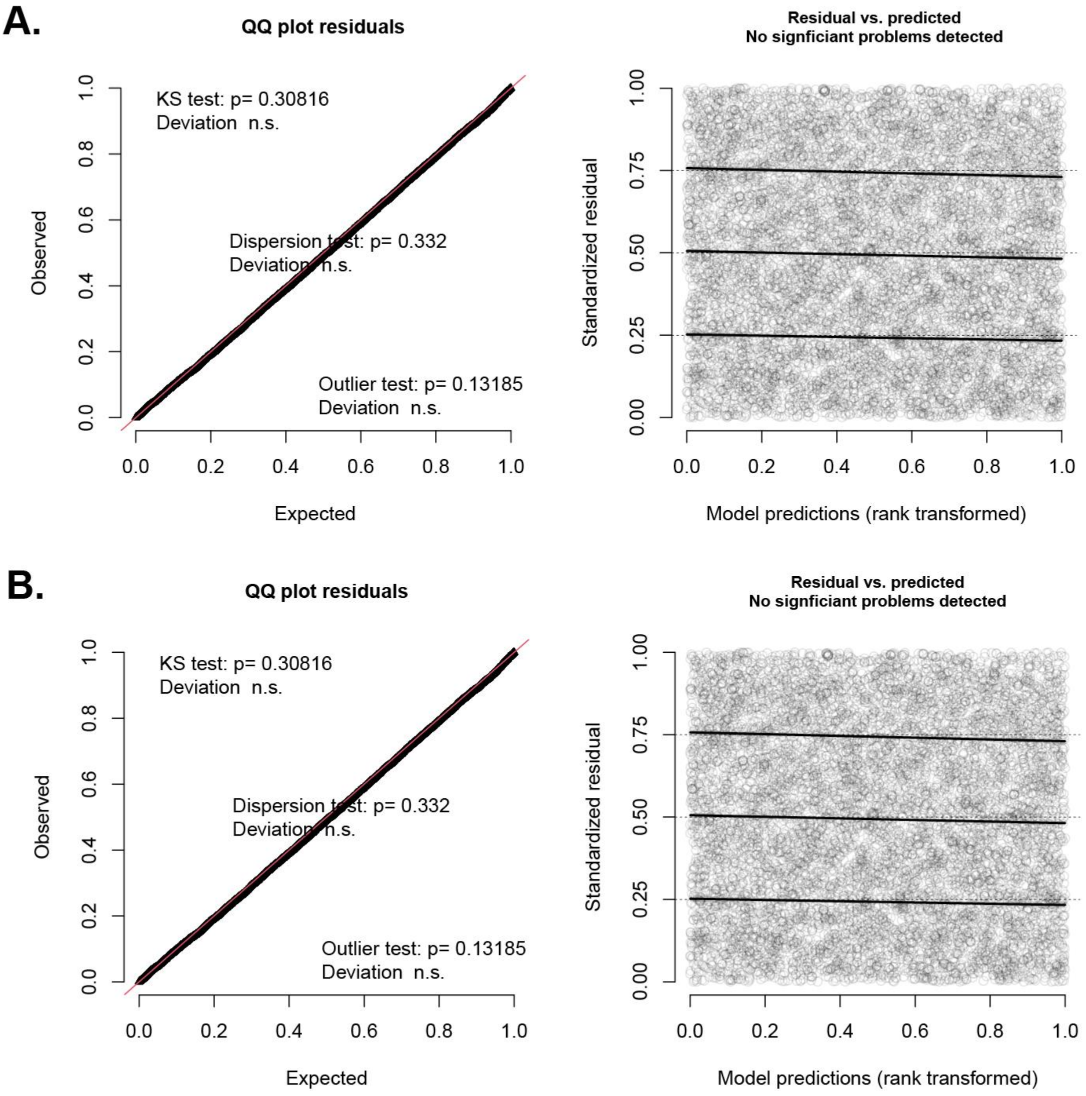

***Figure S**7. DHARMa residual diagnostics for GLMM models. A: DHARMa residual diagnostics for model 1. B: DHARMa residual diagnostics for model 2.*

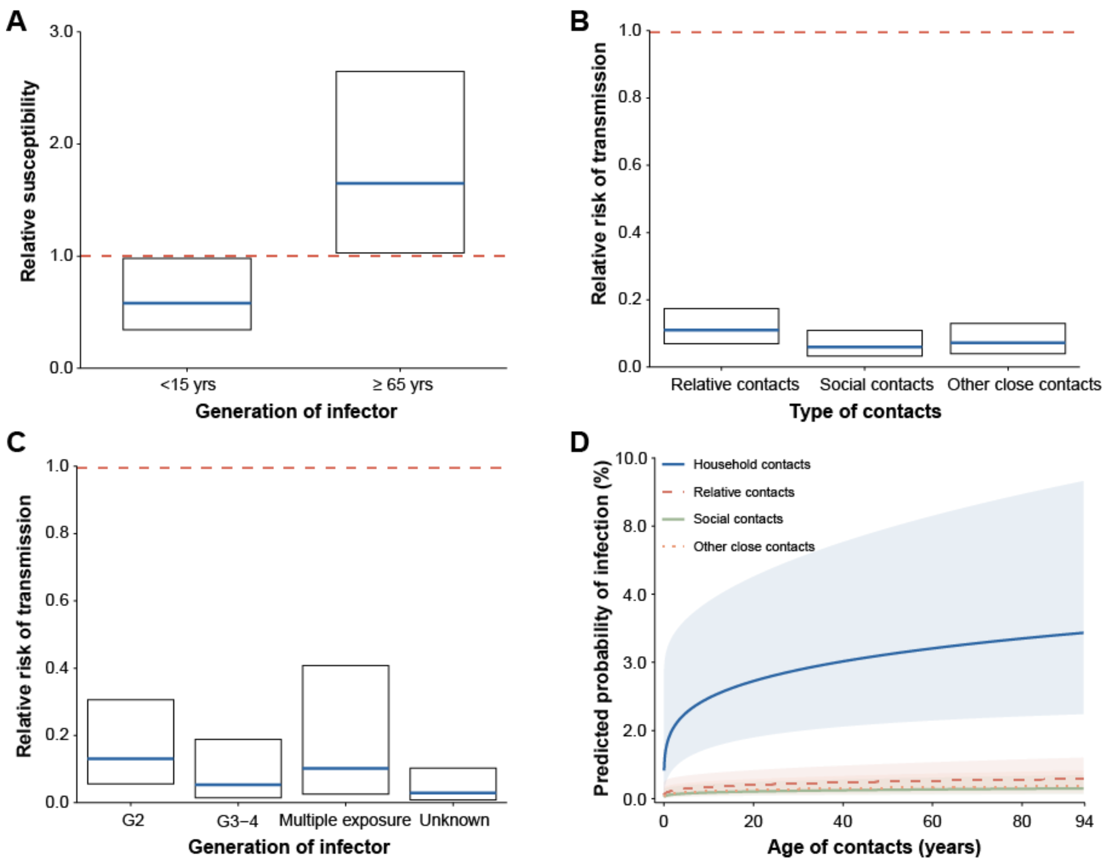

***Figure S8.*** *Potential drivers of COVID-19 transmissions in Hunan Province, China. (A) The relative susceptibility of contacts who were younger than 15 years of age and of those who were older than 65 years of age (the reference group [red lines] was contacts who were 15-64 years of age); (B) The relative risk of SARS-CoV-2 infections among contacts with different type of exposures to an infector (the reference group [red lines] was contacts who were exposed to an infector in households); (C) The relative risk of SARS-CoV-2 infections among contacts with exposures to infectors with different generations of SARS-CoV-2 transmission (the reference group [red lines] was contacts who were exposed to an index case-patients); (D) The probability of SARS-CoV-2 infections at a given age of contacts in a specific setting.*

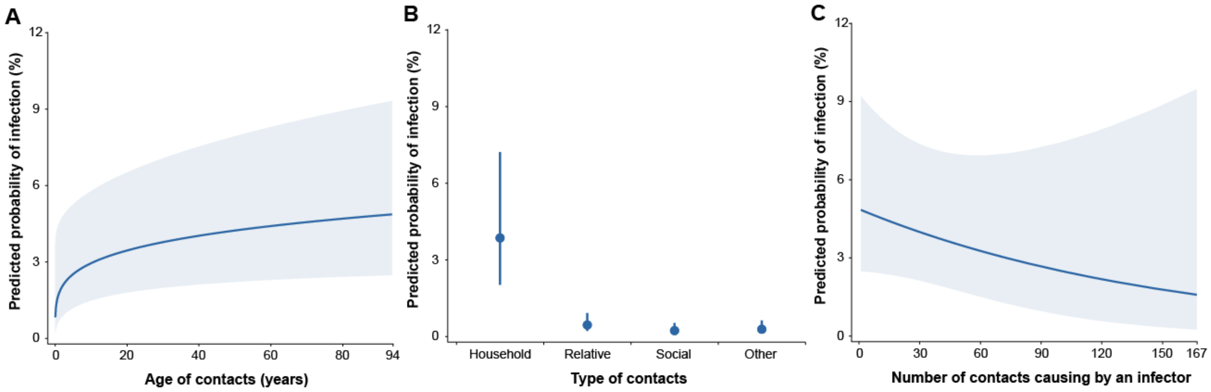

***Figure S9.*** *The marginal effects of various covariates in GLMM model. The smoothed lines and shared areas represent the point estimates and 95% confidence interval for the probability of SARS-CoV-2 infections, respectively.*

***Table S13.*** *Stepwise regression analysis of factors associated with the probability of acquiring SARS-CoV-2 infections in generalized additive mixed models with splines for age of infectors and contacts, as well as number of contacts.*

| Characteristics | No. of contact | Step 4-1 | | Step 4-2 | |
|---|---|---|---|---|---|
| | | OR (95%CI) | P-value | OR (95%CI) | P-value |
| **Parametric coefficients** | | | | | |
| Intercept | - | 0.25 (0.12, 0.51) | <0.001 | 0.23 (0.11, 0.47) | <0.001 |
| Age of infectors | | | | | |
| 0-14 y | 193 | 0.33 (0.06, 1.66) | 0.177 | - | - |
| 15-64 y | 6,833 | Reference | - | - | - |
| 65+ y | 1,133 | 0.60 (0.29, 1.23) | 0.166 | - | - |
| Age of contacts | | | | | |
| 0-14 y | 936 | 0.62 (0.38, 1.01) | 0.055 | - | - |
| 15-64 y | 6,411 | Reference | - | - | - |
| 65+ y | 812 | 1.54 (0.99, 2.39) | 0.055 | - | - |
| Type of contact | | | | | |
| Household contacts | 1,021 | Reference | - | Reference | - |
| Relative contacts | 3,084 | 0.14 (0.09, 0.21) | <0.001 | 0.14 (0.09, 0.21) | <0.001 |
| Social contacts | 2,227 | 0.09 (0.05, 0.15) | <0.001 | 0.08 (0.05, 0.14) | <0.001 |
| Other contacts | 1,827 | 0.10 (0.06, 0.17) | <0.001 | 0.10 (0.06, 0.17) | <0.001 |
| Generation of SARS-CoV-2 transmission | | | | | |
| G1 | 2,121 | Reference | - | Reference | - |
| G2 | 2,987 | 0.27 (0.15, 0.48) | <0.001 | 0.27 (0.15, 0.48) | <0.001 |
| G3-4 | 965 | 0.17 (0.08, 0.38) | <0.001 | 0.18 (0.08, 0.40) | <0.001 |
| Multiple exposure[b] | 598 | 0.26 (0.10, 0.70) | 0.007 | 0.28 (0.10, 0.75) | 0.011 |
| Unknown | 1,488 | 0.07 (0.03, 0.18) | <0.001 | 0.08 (0.03, 0.20) | <0.001 |
| Gender of infectors | | | | | |
| Female | 4,067 | Reference | - | Reference | - |
| Male | 4,092 | 1.62 (1.02, 2.57) | 0.041 | 1.65 (1.04, 2.62) | 0.035 |
| Gender of contacts | | | | | |
| Female | 4,017 | Reference | - | Reference | - |

| | | | | | |
|---|---|---|---|---|---|
| Male | 4,142 | 1.01 (0.74, 1.36) | 0.963 | 1.00 (0.74, 1.35) | 0.988 |
| **Approximate significance of smooth terms** | | | | | |
| Random effect | | | | | |
| Infector | - | - | <0.001 | - | <0.001 |
| Cluster | - | - | 0.141 | - | 0.124 |
| Age of infectors | | | | | |
| Spline for age | - | - | - | - | 0.198 |
| Age of contacts | | | | | |
| Spline for age | - | - | - | - | <0.001 |
| Levels of exposure to an infector | | | | | |
| Spline for number of contacts | - | - | 0.032 | - | 0.040 |

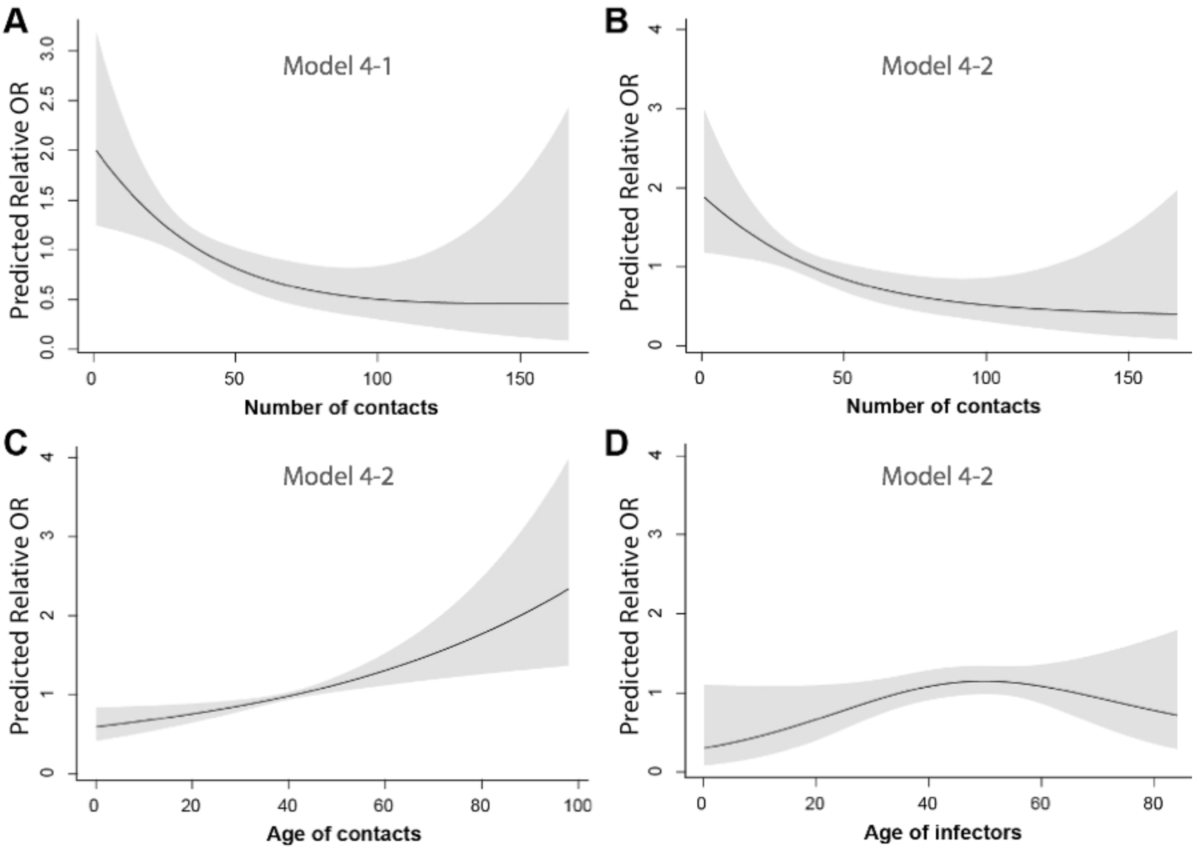

***Figure S10.*** *GAMM-predicted non-linear 3-knot splines for age of infector, age of contacts, and the number of contacts. A. model 4-1; B to D. model 4-2. The shaded area delimits the 95% confidence intervals of the spline functions.*

### References

1. National Health Commission of the People's Republic of China. Diagnosis and treatment guideline on pneumonia infection with 2019 novel coronavirus (6th trial edn). 2020.

2. He X, Lau EHY, Wu P, et al. Temporal dynamics in viral shedding and transmissibility of COVID-19. *Nat Med* 2020; **26**(5): 672-5.

3. Ashcroft P, Huisman JS, Lehtinen S, et al. COVID-19 infectivity pro1le correction. *ArXiv e-prints* 2020.

4. Florian Hartig. DHARMa: Residual Diagnostics for Hierarchical (Multi-Level / Mixed) Regression Models. 2016.
